# Effect of Social Media Constraints on Mental Health: A Systematic Review and Meta-Analysis of Experiments

**DOI:** 10.64898/2026.06.01.26354614

**Authors:** Marcus V. V. Lopes, Karina Branje, Athourina David, Anamarie Gennara, Jonathan Haidt, Zachary Rausch, Nikolaus Greb, Anum Aslam, Jacob Lebwohl, Jean-Philippe Chaput, Gary S. Goldfield

## Abstract

**Background:** Observational studies have consistently reported associations between social media use (SMU) and poorer mental health outcomes; however, such designs cannot establish causality. This study synthesised evidence from randomized experiments to estimate the effects of restricting SMU on mental health outcomes.

**Methods:** A systematic search was conducted across MEDLINE, Embase, PsycINFO, and Cochrane CENTRAL to identify experimental trials evaluating interventions that constrained SMU for at least 24 hours and included an unconstrained control condition. Multilevel random-effects meta-analyses were used to synthesise effect estimates. Prespecified meta-regressions explored study-level moderators, and population-level impact fractions were estimated relative to global SMU prevalence.

**Results:** From 7,784 screened records, 37 reports representing 35 distinct studies were included (pooled N = 7,160). Most interventions lasted one to three weeks and targeted college-aged youth. Pooled estimates favoured SMU constraints across outcomes, with magnitude and precision varying by domain. Confidence intervals were entirely above zero, consistent with a beneficial response for depressive symptoms (g = 0.22; 95% CI, 0.12 to 0.32), perceived stress (g = 0.15; 95% CI, 0.01 to 0.29), anxiety symptoms (g = 0.19; 95% CI, 0.05 to 0.34), fear of missing out/nomophobia (g = 0.14; 95% CI, 0.04 to 0.24), well-being (g = 0.36; 95% CI, 0.10 to 0.63) and body image (g = 0.26; 95% CI, 0.02 to 0.50). Heterogeneity was substantial for several outcomes (I^2^ > 75%). In bivariate meta-regressions, higher baseline SMU was associated with larger effects for anxiety symptoms (β = 0.13; 95% CI, 0.03 to 0.22), and longer interventions were associated with larger effects for depressive symptoms (β = 0.16; 95% CI, 0.02 to 0.30). Inferences revealed that a short-term reduction in SMU globally could plausibly mitigate 17.5% and 15.4% of depressive and anxiety symptom cases, respectively.

**Conclusions:** Experimental design-based evidence supports the causal case for an effect of SMU on mental health, with constraints producing improvements across multiple outcomes and no evidence of harm. Population-level inferences suggest that even individually modest effects may translate into meaningful public health benefits given the high prevalence of SMU exposure. These findings suggest that reducing SMU may represent a low-intensity, low-cost, scalable strategy to support mental health and improve well-being.

## Introduction

Mental health disorders are a leading cause of disability worldwide, with anxiety and depressive disorders alone affecting an estimated 359 and 332 million people, respectively, together accounting for 63% of all mental health disorders.^1^ In parallel with the increasing burden of mental distress, social media use (SMU) has become pervasive across age groups: 77% of US high school students report using social media at least several times a day,^2^ approximately half of US adults visit Facebook or YouTube daily,^3^ and 43% of youth aged 11–15 across 40 countries report intense SMU.^4^ Although social media has been proposed as a tool to foster social connections, platform design features developed over the past decade may exploit developmental vulnerabilities,^5^ potentially promoting excessive use and amplifying psychological harm during adolescence and early adulthood, a period marked by heightened risk-taking, sensitivity to social rejection, emotional reactivity, and identity formation.

The potential link between social media usage and negative developmental outcomes is a growing concern among parents, educators, clinicians, researchers and policy makers and is supported by accumulating empirical evidence. Meta-analyses of observational studies have consistently linked higher SMU to greater depressive and anxiety symptoms,^6–8^ poorer subjective well-being,^9–11^ and more negative self-evaluations.^12^ However, observational designs cannot establish whether these associations are causal, motivating a growing number of randomized experiments examining whether constraining SMU improves mental health.

Previous evidence syntheses of these trials generally conclude on inconclusive findings or modest mental health benefits of constraining social media.^13–20^ However, most syntheses aggregated effects across heterogeneous psychological constructs which mask domain-specific effects and hinder clinical interpretation.^14–16,18^ Although original experimental studies usually applied appropriate analyses for their respective designs, none of the previous evidence syntheses incorporated pre-post correlations for variance estimation, which can distort precision and alter study weights. Finally, prior reviews have relied on conventional effect-size benchmarks to interpret findings, often dismissing pooled estimates as trivially small without considering their population-level implications.

To address these limitations and gain a clearer understanding of the causal impacts of SMU on mental health, we conducted a systematic review and meta-analysis of randomized controlled trials (RCT) to address the following study objectives: 1) to estimate the effects of constraining social media use on domain-specific mental health outcomes; 2) to examine whether the effects vary by study-level characteristics; and 3) to provide plausible estimates of the potential public health impact of reducing SMU on mental health.

## Methods

### Search strategy and selection criteria

This review was registered with the International Prospective Register of Systematic Reviews (PROSPERO; CRD42025637996) and was conducted in accordance with the Preferred Reporting Items for Systematic Reviews and Meta-Analyses (PRISMA 2020) statement (Appendix p 2-4).^23^ Deviations from the registered protocol, along with their rationale, are documented in the appendix (p 5-6).

We included RCTs employing between-group or mixed designs examining the effect of social media constraints on mental health outcomes. Eligible studies enrolled participants of any age and clinical status. Interventions were required to constrain SMU for a minimum duration of 24 hours via daily time limits, relative reductions, or complete abstinence compared with an unmanipulated behavioral control group. Studies that incorporated additional mental health-promoting components beyond social media constraint (e.g., cognitive behavioural therapy) were excluded to isolate the independent effect of SMU constraints. Operational definitions for social media and mental health outcomes are provided in registered protocol.

Electronic searches were conducted in MEDLINE, Embase, APA PsycINFO, and Cochrane CENTRAL via the Ovid platform on November 19, 2024, and then updated on October 3, 2025. No language restriction was applied. Reference lists of included studies and relevant reviews were also screened to identify additional eligible trials. The full electronic search strategies for all databases are provided in the appendix (p 7-9).

Following duplicate removal, records were imported into Covidence (Veritas Health Innovation, Melbourne, Australia) for eligibility screening. Title and abstract screening, full-text review, data extraction and risk of bias assessments (Cochrane Risk of Bias 2 tool) were performed independently and in duplicate by two reviewers. Disagreements at any stage were resolved by MVVL. Corresponding authors were contacted to obtain missing summary statistics. In cases where individual-participant data was available, summary statistics aggregated at the study-level were re-estimated to ensure precision. Details on quality assurance procedures are described in appendix (p 10).

### Effect Measures

Effect sizes were calculated from study-level means and standard deviations (SD) of outcomes per condition and were expressed as standardized mean differences (SMDs) with correction for small samples (Hedges’s g). For studies utilizing a pre-test/post-test control group design, we calculated SMDs and their corresponding sampling variances using the Morris (2008) *d*_*ppc2*_ estimator which incorporates pre-post correlation coefficient (*r*)^22^ When individual participant data were publicly available, study-specific pre-post correlations were calculated empirically for each outcome. Missing pre-post correlation values were imputed using domain-specific averages (appendix p 13) or set to a conservative fallback of *r* = 0 60 (i.e., rounded from the lowest *r* among averages). For studies implementing a between-groups design at post-intervention only, SMD was computed using the pooled post-test SD. Where summary statistics were insufficient, effect sizes were derived from F-statistics via standard conversion formulae.^23^ Effect signs were reversed for outcomes where lower scores indicate clinical improvement, so that positive estimates uniformly reflect beneficial effects of SMU constraints. Full computational details and R implementation are provided in the appendix (appendix p 11-16).

### Data Synthesis

#### Meta-analysis

Multilevel random-effects models were fitted using restricted maximum likelihood (REML) estimation. To account for statistical dependency arising from multiple samples or intervention arms contributed by the same study, random intercepts were specified at both the study level and the sample level, with samples nested within studies.

Statistical inference was based on t-distribution-based tests with containment degrees of freedom. Between-study heterogeneity was quantified using the I^2^ statistic adapted for multilevel models and evaluated using Cochran’s Q-type omnibus test. Small-study effects were examined through visual inspection of funnel plots and assessed using an Egger-type regression test when at least 10 effect estimates were available for a given outcome (k ≥10). The influence of individual studies on pooled estimates was evaluated using leave-one-out sensitivity analyses.

The analyses were performed in R, version 4.5.1 for Windows (R Foundation for Statistical Computing, Vienna, Austria), using the *metafor* package. Interpretation relied on the magnitude and precision of effect estimates, with confidence intervals and p-values used as continuous indicators of compatibility with a range of plausible effects.^24–26^

#### Study-level moderators

Sources of heterogeneity were explored using meta-regression within the multilevel random-effects framework. Prespecified study-level characteristics were examined as moderators of intervention effects, including region (based on World Bank definitions), study participants’ mean age, gender/sex distribution (expressed as proportion of females/women), SMU at baseline, sample type (whether participants were drawn from the general population or symptomatic groups), intervention length, type of social media constraint (reduction vs abstinence), year of data collection, and risk-of-bias level. Moderator effects were estimated by including fixed-effect terms in the multilevel models, while retaining random intercepts at the study and sample levels to account for within-study dependency. Other planned moderators were not tested due to reporting inconsistency (see Appendix, p 7-8). Given that moderator analyses in meta-analytic frameworks are inherently exploratory and often underpowered,^27^ these analyses were conducted only for outcomes with at least 10 independent samples, consistent with Cochrane guidance.^28^

### Sensitivity analysis

A series of sensitivity analyses were conducted to evaluate the robustness of the pooled estimates to key methodological assumptions. First, robustness to effect size specification was examined by recalculating pooled effects using alternative estimators, including d_PPC1_^27^ (which allows group-specific baseline standardization) and post-intervention-only SMDs, the latter replicating approaches used in prior meta-analyses.^13^ Second, analyses were repeated after excluding effect estimates derived from studies classified as potentially problematic. These included studies with (1) outlying or highly influential effect sizes, (2) indications of small-sample bias, (3) evidence of unsuccessful intervention implementation (e.g., non-adherence to social media reduction or abstinence protocols), or (4) control groups in which absence of manipulation was not clearly confirmed. Third, analyses excluding outcomes measured using single-item indicators rather than validated multi-item scales were performed, given the greater susceptibility of single-item measures to measurement error and random variability, which may artificially inflate standardized effect sizes. Fourth, analyses excluding between-group RCTs that relied solely on post-intervention comparisons were conducted, as such designs do not permit estimation of change-score-based effect sizes (i.e., d_PPC1_ and d_PPC2_).^27^

#### Population-level inferences

To move beyond the interpretation of effect sizes based on arbitrary magnitude benchmarks and instead situate findings within meaningful real-world contexts,^29^ we estimated the potential public health impact of reducing SMU on mental health by computing the Potential Impact Fraction (PIF) for each mental health outcome within the internalizing symptoms domain (i.e., depressive and anxiety symptoms). The PIF quantifies the proportion of cases potentially preventable under a specified counterfactual reduction in exposure prevalence, ranging from partial reduction to complete elimination^30^. For each outcome, the meta-analytic effect size was converted to an odds ratio using the Cox method,^23^ and used as an approximation of the relative risk given the absence of outcome prevalence data among unexposed youth. These relative risks were combined with the observed prevalence of high SMU among youth – operationalized as the combined proportion of intense users (having online contact with others almost all the time throughout the day with non-problematic SMU) and problematic users (endorsing six or more symptoms of problematic use regardless of online contact frequency) survey across 40 countries (HBSC 2021/2022),^4^ to estimate the PIF under multiple counterfactual scenarios. High SMU was selected as a conservative exposure definition rather than any use, as it more closely reflects the exposure profile of participants included in the reviewed randomized clinical trials.

## Results

### Study Selection

The database search identified 7,784 records, and an additional 17 records were identified through citation searching (Figure 1). After removal of 2,372 duplicates, 5,412 records were screened at the title/abstract level, and 5,349 were excluded. Full texts were sought for 63 reports from databases and 17 reports from other methods; all were retrieved and assessed for eligibility. Among reports assessed for eligibility, 35 studies reported across 37 reports met all criteria and were included in the review and meta-analysis. Reasons for exclusion are reported in appendix (p 17-20).

**Figure 1.**
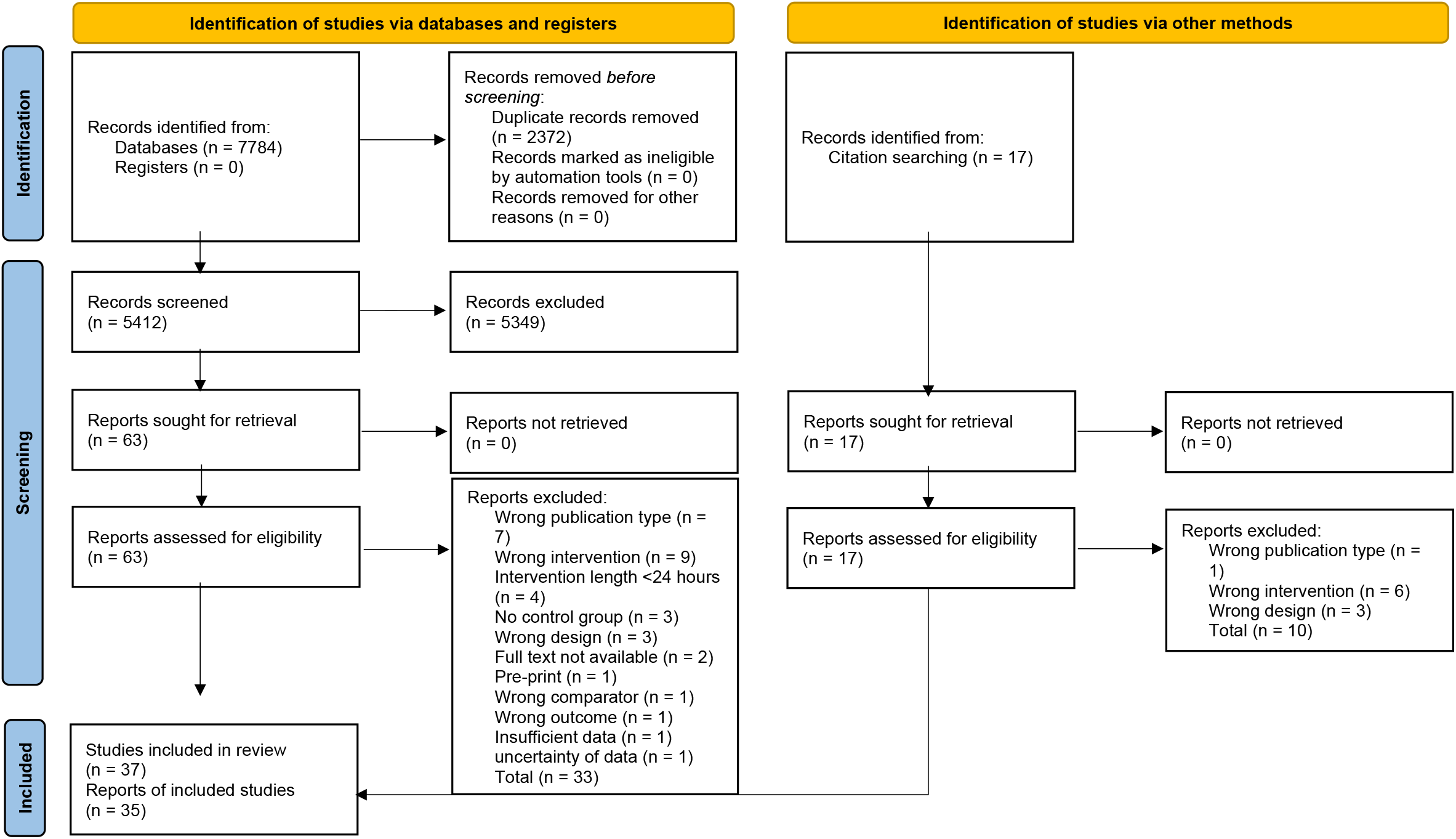
PRISMA 2020 Flow Diagram of Study Selection

### Study Characteristics

Study characteristics are presented in appendix (p 21-30). Most studies were conducted in Europe (14/35; 40%) and in North America (13/35; 37%). Samples were predominantly drawn from the general population (32/35; 91%), with fewer screened for symptoms of distress (3/35; 9%). Interventions more commonly used SMU abstinence designs (20/35; 57%) than SMU reduction designs (15/35; 43%) for an average duration of 1.88 ± 1.75 weeks. Data were collected between 2015-2024 (median = 2020). Risk of bias was rated as low risk for 2/35 unique studies (6%), some concerns for 26/35 (74%), and high risk for 7/35 (20%). Details on risk of bias assessment are reported in appendix (p 31-32). Participants across studies were mostly youth (mean age: 27.3 ± 5.1 years; sex: 68% females) who spent an average of 99 ± 41.6 minutes/day on social media.

### Study-level meta-analytical effects

The summary of meta-analyses by outcome is presented in Table 1. The pooled effects of social media constraints on mental health were generally in the positive direction (i.e., beneficial), although the magnitude and precision varied by outcome. Point estimates for depressive symptoms (g=0.22; 95% CI, 0.12 to 0.32), perceived stress (g=0.15; 95% CI, 0.01 to 0.29), anxiety symptoms (g=0.19; 95% CI, 0.05 to 0.34), fear of missing out (FoMO)/nomophobia (g=0.14; 95% CI, 0.04 to 0.24), well-being (g=0.36; 95% CI, 0.10 to 0.63) and body image (g = 0.26; 95% CI, 0.02 to 0.50) were accompanied by confidence intervals situated entirely above zero, supporting a beneficial direction of effect across each of these outcomes.

**Table 1.** Pooled meta-analytic estimates by outcome.

| Outcome | Main Effects |  |  | Heterogeneity |  |  | Publication Bias |
| --- | --- | --- | --- | --- | --- | --- | --- |
|  | *k* | N | Hedges' *g* [95% CI] | *Q* | *pQ* | *I*<sup>2</sup> (%) | Egger's *p* |
| **Total studies** |  |  |  |  |  |  |  |
| Depressive Symptoms | 18 (18) | 4335 | **0.22 [0.12, 0.32]** | 54.964 | 0 | 66.15 | 0.011 |
| Perceived Stress | 7 (7) | 1169 | **0.15 [0.01, 0.29]** | 4.779 | 0.572 | 0 | NA |
| Anxiety Symptoms | 11 (11) | 2969 | **0.19 [0.05, 0.34]** | 30.656 | 0.001 | 67.32 | 0.5 |
| Loneliness/Isolation | 14 (18) | 3259 | 0.09 [-0.01, 0.20] | 41.731 | 0.001 | 62.24 | 0.506 |
| FoMO/Nomophobia | 9 (10) | 1214 | **0.14 [0.04, 0.24]** | 5.417 | 0.797 | 0 | 0.411 |
| Negative Affect/Emotions | 8 (8) | 876 | 0.07 [-0.29, 0.43] | 61.872 | 0 | 89.04 | NA |
| Well-being | 11 (12) | 3805 | **0.36 [0.10, 0.63]** | 92.208 | 0 | 92.9 | 0.379 |
| Self-Esteem | 7 (9) | 718 | 0.12 [-0.16, 0.40] | 74.214 | 0 | 87.97 | NA |
| Happiness/Positive Affect | 13 (14) | 3427 | 0.12 [-0.02, 0.25] | 33.133 | 0.002 | 67.1 | 0.244 |
| Life Satisfaction | 14 (16) | 4419 | 0.11 [-0.03, 0.24] | 41.94 | 0 | 75.34 | 0.57 |
| Body Image | 6 (18) | 569 | **0.26 [0.02, 0.50]** | 68.242 | 0.000 | 80.26 | 0.581 |
| Eating Disorders/Pathology | 2 (2) | 244 | 0.15 [-3.01, 3.31] | 6.898 | 0.009 | 85.5 | NA |
| **Problematic cases removed** |  |  |  |  |  |  |  |
| Depressive Symptoms | 16 (16) | 2663 | **0.21 [0.11, 0.30]** | 28.122 | 0.021 | 46.84 | 0.68 |
| Anxiety Symptoms | 9 (9) | 1231 | **0.27 [0.14, 0.39]** | 11.966 | 0.153 | 30.11 | NA |
| Loneliness/Isolation | 12 (16) | 1542 | **0.13 [0.03, 0.23]** | 30.035 | 0.012 | 44.9 | 0.098 |
| FoMO/Nomophobia | 8 (9) | 1138 | **0.15 [0.04, 0.26]** | 4.261 | 0.833 | 0 | NA |
| Negative Affect/Emotions | 7 (7) | 800 | 0.03 [-0.39, 0.45] | 60.294 | 0 | 90.47 | NA |
| Well-being | 10 (10) | 2148 | **0.30 [0.08, 0.52]** | 42.662 | 0 | 82.7 | 0.854 |
| Happiness/Positive Affect | 10 (11) | 1632 | 0.08 [-0.05, 0.22] | 20.769 | 0.023 | 45.89 | 0.507 |
| Life Satisfaction | 12 (14) | 2656 | **0.15 [0.03, 0.26]** | 25.549 | 0.02 | 51.99 | 0.415 |
| Body Image | 6 (18) | 569 | **0.26 [0.02, 0.50]** | 68.242 | 0.000 | 80.26 | 0.581 |

Note: *k*: Number of studies [number of effect sizes]. *N*: Total sample size. Hedges' *g*: Pooled effect size. *QE*: Q-statistic for heterogeneity. *QEp*: p-value for Q-statistic. *I*<sup>2</sup>:
Percentage of total variability due to true effect size differences. Egger's *p*: p-value for Egger's regression test for publication bias. FoMO: fear of missing out. NA: Not available due to insufficient studies for testing.

Heterogeneity was substantial for several outcomes (I^2^ >75% for negative affect/emotions, well-being; self-esteem; life satisfaction; and body image), suggesting that intervention effects varied across studies and contexts. Two studies were flagged as potentially problematic by design or implementation: Van Wezel et al. (2021) reported unsuccessful experimental manipulation, and Allcott et al. (2020) reported brief behavioral manipulation in the control condition, which limits comparability with trials using unmanipulated controls.For depressive symptoms, the Egger test indicated possible small-study bias (p=.011) – a pattern in which smaller studies report disproportionately larger effects – attributable largely to Reed et al. (2023), which reported an exceptionally large effect size. Leave-one-out analyses further indicated that heterogeneity was partially attributable to a small number of outlying or influential studies. Details on studies flagged as problematic by outcome are provided in appendix (p 33).

Sensitivity analyses excluding problematic cases by design or implementation and outlying effects led to considerable changes on pooled effect sizes for anxiety symptoms (g=0.27; 95% CI, 0.14 to 0.39), loneliness (g=0.13; 95% CI, 0.03 to 0.23), well-being (g=0.30; 95% CI, 0.08 to 0.52), and life satisfaction (g=0.15; 95% CI, 0.03 to 0.26). Excluding outcomes measured with single-item indicators yielded slightly larger pooled effects than the main analyses. Re-analyses using alternative effect-size estimators showed that d_PPC1_ and d_PPC2_-derived effects were highly similar. In contrast, post-intervention-only SMDs generally yielded less precise estimates (wider confidence intervals) and, for several outcomes, smaller pooled effects than d_PPC2_.

All sensitivity analyses are reported in the appendix (p 34-39). Funnel plots and leave-one-out analyses are available for inspection at the interactive web-based application.

### Study-level moderation effects

Bivariate meta-regressions suggested outcome-specific moderation (Table 2). Higher baseline SMU was associated with larger effects for anxiety symptoms (β=0.13; 95% CI 0.03 to 0.22), and longer interventions had larger effects for depressive symptoms (β=0.16; 95% CI 0.02 to 0.30). For loneliness/isolation, effects were larger in SMU reduction experiments (β=0.24; 95% CI 0.08 to 0.39) than abstinence interventions (β=0.04; 95% CI -0.05 to 0.13). After excluding problematic cases (Appendix, p 37), the intervention-length association attenuated for depressive symptoms (β=0.05; -0.04 to 0.15) but strengthened for loneliness/isolation (β=0.10; 0.01 to 0.20). The associations between SMU levels and the effect for anxiety (β=0.09; -0.05 to 0.22) after problematic cases removal.

**Table 2.** Study-level moderators examined through meta-regression.

| Moderators | Depressive Symptoms |  | Anxiety Symptoms |  | Loneliness/Isolation |  | FoMO/Nomophobia |  | Well-being |  | Happiness/Positive Affect |  | Life Satisfaction |  | Body Image |  |
| --- | --- | --- | --- | --- | --- | --- | --- | --- | --- | --- | --- | --- | --- | --- | --- | --- |
|  | *g* [95% CI] | *p* | *g* [95% CI] | *p* | *g* [95% CI] | *p* | *g* [95% CI] | *p* | *g* [95% CI] | *p* | *g* [95% CI] | *p* | *g* [95% CI] | *p* | *g* [95% CI] | *p* |
| **Region** |  | 0.718 |  | 0.612 |  | 0.733 |  | 0.696 |  | 0.312 |  | 0.44 |  | 0.177 |  | 0.374 |
| East Asia & Pacific | — |  | — |  | — |  | — |  | 0.65 [0.11, 1.18] |  | -0.00 [-0.42, 0.42] |  | -0.16 [-0.46, 0.15] |  | — |  |
| Europe & Central Asia | 0.20 [0.04, 0.37] |  | 0.26 [-0.02, 0.53] |  | 0.16 [-0.03, 0.34] |  | 0.16 [-0.02, 0.35] |  | 0.33 [-0.01, 0.67] |  | 0.23 [-0.01, 0.46] |  | 0.17 [-0.00, 0.33] |  | 0.11 [-0.29, 0.51] |  |
| North America | 0.24 [0.09, 0.38] |  | 0.18 [-0.04, 0.39] |  | 0.12 [-0.00, 0.25] |  | 0.13 [0.00, 0.25] |  | 0.06 [-0.57, 0.69] |  | 0.05 [-0.20, 0.31] |  | 0.09 [-0.14, 0.32] |  | 0.33 [0.00, 0.66] |  |
| **Mean age** | -0.01 [-0.13, 0.11] | 0.879 | 0.04 [-0.15, 0.22] | 0.643 | -0.01 [-0.12, 0.10] | 0.853 | 0.08 [-0.04, 0.21] | 0.143 | -0.02 [-0.30, 0.27] | 0.903 | -0.02 [-0.17, 0.12] | 0.749 | 0.03 [-0.11, 0.17] | 0.636 | -0.14 [-0.32, 0.05] | 0.140 |
| **Proportion of females** | -0.06 [-0.17, 0.06] | 0.337 | -0.02 [-0.18, 0.14] | 0.793 | 0.03 [-0.08, 0.14] | 0.567 | -0.03 [-0.15, 0.08] | 0.534 | 0.14 [-0.13, 0.42] | 0.263 | 0.03 [-0.13, 0.19] | 0.711 | 0.01 [-0.13, 0.16] | 0.833 | 0.03 [-0.15, 0.21] | 0.712 |
| **Social media use at baseline** | 0.04 [-0.10, 0.18] | 0.58 | 0.13 [0.03, 0.22] | 0.015 | 0.01 [-0.16, 0.18] | 0.866 | 0.03 [-0.09, 0.16] | 0.543 | 0.12 [-0.44, 0.67] | 0.593 | 0.08 [-0.07, 0.23] | 0.255 | -0.04 [-0.24, 0.16] | 0.663 | -0.06 [-0.21, 0.10] | 0.329 |
| **Sample type** |  | 0.336 |  | 0.363 |  | — |  | 0.952 |  | — |  | — |  | — |  | 0.543 |
| Clinical sample | 0.34 [0.07, 0.61] |  | 0.34 [-0.03, 0.70] |  | — |  | 0.13 [-0.05, 0.31] |  | — |  | — |  | — |  | 0.17 [-0.19, 0.54] |  |
| Community sample | 0.20 [0.09, 0.31] |  | 0.17 [-0.00, 0.33] |  | — |  | 0.14 [0.02, 0.26] |  | — |  | — |  | — |  | 0.30 [0.05, 0.56] |  |
| **Intervention length (weeks)** | 0.16 [0.02, 0.30] | 0.026 | -0.01 [-0.20, 0.18] | 0.868 | 0.11 [-0.01, 0.23] | 0.062 | -0.04 [-0.14, 0.07] | 0.441 | -0.17 [-0.44, 0.10] | 0.201 | 0.01 [-0.12, 0.14] | 0.875 | -0.02 [-0.16, 0.12] | 0.764 | -0.11 [-0.28, 0.05] | 0.168 |
| **Manipulation type** |  | 0.178 |  | 0.86 |  | 0.03 |  | 0.63 |  | 0.499 |  | 0.884 |  | 0.944 |  | 0.784 |
| Abstinence | 0.15 [0.01, 0.30] |  | 0.18 [-0.05, 0.42] |  | 0.04 [-0.05, 0.13] |  | 0.10 [-0.11, 0.31] |  | 0.29 [-0.05, 0.64] |  | 0.11 [-0.06, 0.29] |  | 0.11 [-0.06, 0.28] |  | 0.25 [0.03, 0.47] |  |
| Reduction | 0.29 [0.14, 0.43] |  | 0.21 [0.00, 0.41] |  | 0.24 [0.08, 0.39] |  | 0.15 [0.04, 0.27] |  | 0.47 [0.04, 0.90] |  | 0.14 [-0.12, 0.39] |  | 0.10 [-0.17, 0.36] |  | 0.27 [0.05, 0.50] |  |
| **Study date** | 0.06 [-0.04, 0.16] | 0.225 | 0.09 [-0.04, 0.22] | 0.154 | 0.02 [-0.09, 0.13] | 0.713 | 0.09 [-0.03, 0.21] | 0.135 | 0.16 [-0.10, 0.43] | 0.194 | 0.08 [-0.06, 0.22] | 0.24 | 0.04 [-0.10, 0.18] | 0.562 | 0.05 [-0.12, 0.22] | 0.520 |
| **Risk of Bias** |  | 0.71 |  | 0.732 |  | 0.154 |  | — |  | 0.942 |  | 0.275 |  | 0.322 |  | — |
| Low risk | 0.12 [-0.21, 0.44] |  | — |  | — |  | — |  | — |  | — |  | — |  | — |  |
| Some concerns | 0.25 [0.11, 0.40] |  | 0.21 [-0.01, 0.44] |  | 0.05 [-0.07, 0.18] |  | — |  | 0.37 [0.03, 0.71] |  | 0.11 [-0.05, 0.27] |  | 0.07 [-0.10, 0.24] |  | — |  |
| High risk | 0.21 [-0.03, 0.45] |  | 0.15 [-0.16, 0.47] |  | 0.22 [0.03, 0.40] |  | — |  | 0.35 [-0.13, 0.83] |  | 0.31 [-0.04, 0.67] |  | 0.28 [-0.12, 0.69] |  | — |  |

Note: *g*: Hedges' *g* (Pooled Effect Size for continuous moderators; Estimated Marginal Mean for categorical levels). *p*: *p*-value (Omnibus Q-test for categorical headers; Slope
coefficient test for continuous variables). CI: Confidence Interval. FoMO: fear of missing out.

Populational inferences to support effects interpretation is presented in Figure 2. Assuming that SMU causally contributes to internalising symptoms, and using a conservative exposure prevalence estimate (43%; HBSC 2021/2022), a 50% population-level reduction in high SMU – from 43% to 21.5% – could plausibly prevent 8.7% (95% CI, 4.8% to 12.5%) of depressive symptom cases and 7.7% (95% CI, 2.6% to 12.5%) of anxiety cases among youth. Under the counterfactual of complete elimination of high SMU, the estimated PIFs were 17.5% (95% CI, 9.4% to 24.8%) for depressive symptoms and 15.4% (95% CI, 4.9% to 24.7%) for anxiety symptoms, representing the upper-bound population-level impact attributable to this exposure under the conservative prevalence definition applied.

**Figure 2.**
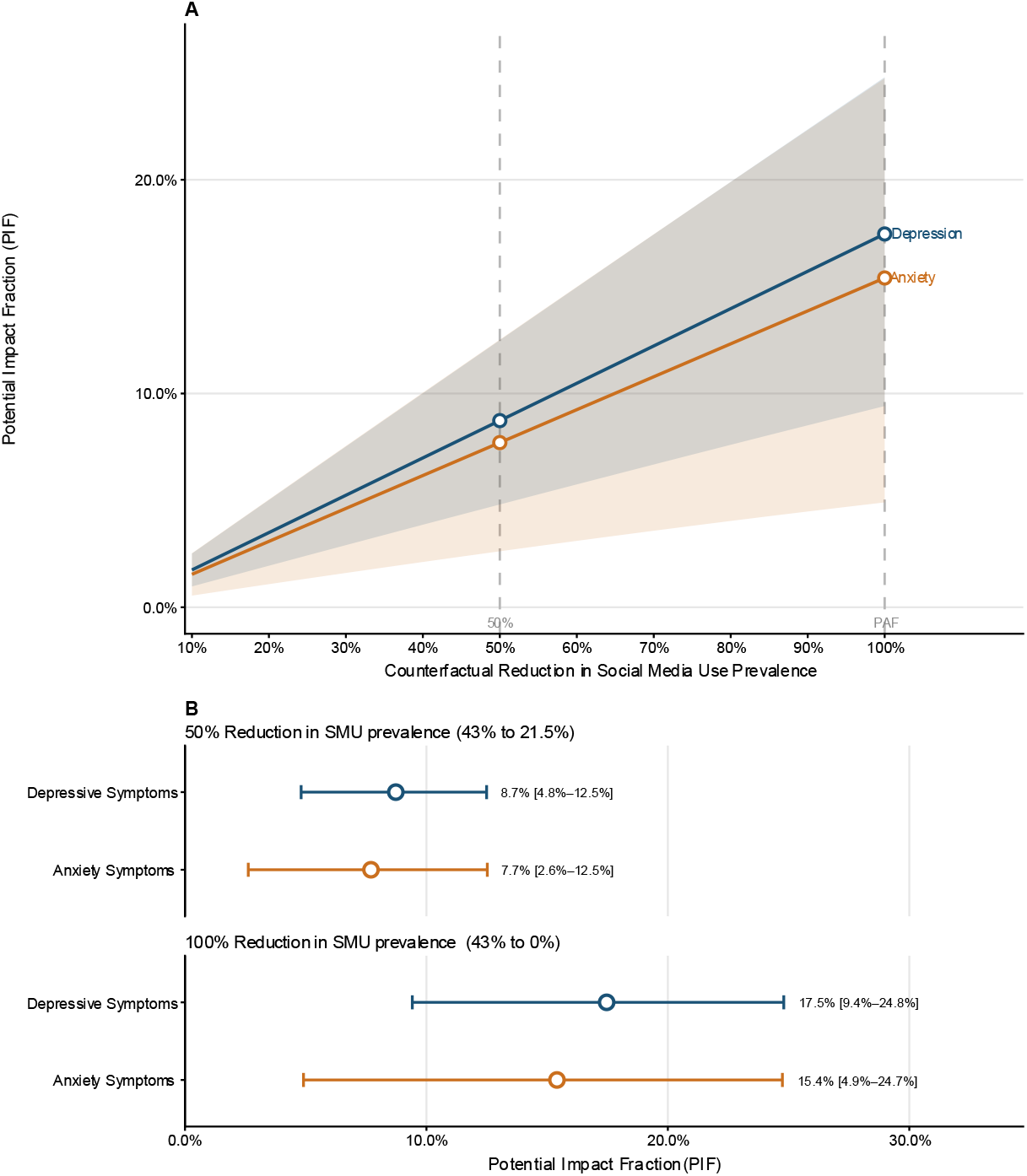
Potential impact fraction of intense social media use on internalising symptoms among youth across counterfactual reduction scenarios. Panel A shows the Potential Impact Fraction (PIF) and 95% confidence intervals across the full range of counterfactual reduction scenarios (10%–100%) for depressive symptoms and anxiety symptoms. Panel B shows PIF point estimates with 95% confidence intervals at two prespecified scenarios: a 50% relative reduction in intense social media use prevalence (from 43% to 21.5%) and complete elimination (100% reduction), which corresponds to the Population Attributable Fraction (PAF). Shaded areas in Panel A and error bars in Panel B represent the 95% confidence intervals.

## Discussion

In this systematic review and meta-analysis of RCTs based on 35 unique studies and 7160 participants, constraining SMU was generally associated with improvements across multiple mental health outcomes. Confidence intervals of point estimates were entirely above zero for internalising symptoms (i.e., depressive and anxiety symptoms) – as well as for perceived stress, FoMO/nomophobia, well-being and body image.

After excluding studies with methodological concerns or influential estimates, beneficial effects were additionally observed for loneliness/isolation and life satisfaction. Conversely, effects for negative and positive affect, self-esteem, and disordered eating were imprecise and broadly compatible with both beneficial or null effects. There was no evidence from point estimates that SMU constraints worsened any outcomes. The imprecise estimates for negative and positive affective outcomes are consistent in direction with the findings of prior meta-analyses restricted to abstinence designs^19^ and those examining a broader range of constraint strategies,^13^ and may partly reflect the composite structure of broad affect scales, which aggregate emotionally distinct states into composite scores and thereby obscure differential effects on specific affective dimensions that symptom-targeted instruments are better positioned to detect.

Our findings extend and attempt to reconcile the inconsistent conclusions of prior experimental syntheses which interpreted the effects of SMU constraints as too small to be practically meaningful,^15^ a conclusion rooted in the application of arbitrary effect size benchmarks that ignore the prevalence of the exposure and the clinical context in which the effect operates.^34^ Conservative population-level inferences indicate that the projected impact of constraining SMU for approximately two weeks is not trivial. Given the high prevalence of intense SMU among youth, our estimates suggest that approximately 17.5% and 15.4% of depressive and anxiety symptom cases, respectively, could theoretically be prevented if intense use were eliminated entirely. Considering that depressive and anxiety disorders are among the leading causes of years lived with disability, collectively affecting over 690 million people worldwide in 2021,^1^ even individually modest effects may translate into meaningful public health benefits. The present findings are broadly consistent in direction and magnitude with those reported by Burnell et al.,^13^ May et al.,^17^ and Lemahieu et al.,^19^ and strengthen the overall causal evidence base by analysing outcomes separately by construct and incorporating empirically derived pre-post correlations into effect size estimation.

Moderation analyses indicated that sources of heterogeneity were outcome specific. Higher baseline SMU was associated with larger effects for anxiety symptoms, suggesting that individuals with heavier use may derive greater benefit from constraint; however, this association attenuated after excluding potentially problematic studies, indicating it may be partly driven by influential cases. Longer intervention duration was associated with larger effects for depressive symptoms in the main analysis, though this association similarly attenuated in sensitivity analyses. For loneliness/isolation, reduction-based interventions were associated with larger effects than abstinence designs, which may reflect that gradual reductions are easier to sustain and better preserve beneficial social contact whilst reducing maladaptive engagement patterns. Meta-regressions were based on study-level aggregates and cannot rule out ecological confounding. Subgroup effects in these domains require further examination using individual participant data meta-analysis.

This review addresses recurring methodological limitations in prior experimental syntheses. Most previous meta-analyses aggregated outcomes across distinct psychological constructs,^14–16^ which risks masking domain-specific effects. Analysing outcomes separately within a multilevel random-effects framework enabled detection of effects that would otherwise be diluted when constructs are pooled. The Morris d_PPC2_ estimator was used as the primary effect size metric, incorporating empirically derived pre-post correlations into the estimation of sampling variances, thereby improving precision estimates relative to approaches that assume independence between baseline and follow-up measurements. This approach reduces sensitivity to baseline imbalance and avoids standardizing effects using postintervention variability that may itself be influenced by the intervention. Sensitivity analyses confirmed that pooled estimates from d_PPC2_ and d_PPC1_ were highly similar, whereas post-intervention-only SMDs yielded less consistent estimates across outcomes, inflating effects for FoMO/nomophobia and self-esteem while attenuating effects for life satisfaction, well-being, and body image. Finally, the review was conducted with an open, reproducible workflow and accompanied by an interactive application that facilitates transparent inspection of pooled results and study-level influences.

Several limitations should be considered when interpreting these findings. The available evidence base is dominated by youth, predominantly female university student samples, which limits generalisability to older adults, youth outside academic settings, and clinical populations. The intervention durations were relatively short on average, and the extent to which the observed mental health effects persist over the long term remains uncertain. Although most studies were rated as having only “some concerns” for risk of bias, a sizable minority (20%) were rated as high risk despite their experimental designs, underscoring the need for greater methodological rigour in future trials. Moderator analyses were conducted at the study level and are therefore vulnerable to ecological bias; observed associations may reflect unmeasured study characteristics rather than genuine individual-level effects and should be treated as hypothesis-generating. Our population-level inferences are limited by the extent of the reviewed RCTs and the representativeness of the epidemiological source used to derive population-level estimates. As such, they should be cautiously interpreted as approximate, plausible estimates. Future research should examine whether partial reductions combined with behavioural substitution – such as structured social or physical activities – produce larger or more durable benefits than constraints alone, using factorial or dismantling designs that permit attribution of effects to specific intervention components. Individual participant data meta-analyses would allow examination of genuine individual-level moderation by baseline symptom severity, usage patterns, and demographic characteristics that cannot be ascertained from study-level aggregates.

### Conclusions

Experimental design-based evidence indicates improvements in multiple mental health outcomes following SMU constraints. No evidence was found that SMU constraints were associated with worsening of any mental health outcome. Population-level inferences suggest that even individually modest effects may translate into meaningful public health benefits given the high prevalence of SMU exposure. These findings support considering reductions in SMU as a feasible, low-intensity strategy for mental health promotion and psychiatric disorder prevention, while recognising that effects were heterogeneous and not consistently observed for all outcomes. Given the scalability and low cost of SMU constraints, these findings lend support to population-level policy initiatives, including school-based social media restrictions, age-based platform regulations, and government legislation requiring transparency and design-level safeguards against excessive use. At the clinical level, SMU reduction may serve as a low-burden adjunct to existing evidence-based interventions, with the potential for complementary behavioural components to enhance and sustain treatment outcomes.

## Data Availability

All analyses conducted in this study are fully reproducible. The study-level data will be made publicly available on the Open Science Framework (OSF) shortly following the publication of this pre-print version and will be accessible at https://osf.io/r37mu. Concurrently, the complete analytic codebase used the meta-analysis, and the interactive web application will be released in a single GitHub repository https://github.com/MarcusVVLopes/smu_meta_repo. The live application, deployed at https://marcuslopes.shinyapps.io/smu_meta/, allows for real-time visualization of study-level and pooled estimates under alternative sensitivity analysis criteria to enhance transparency and facilitate knowledge translation.

## Acknowledgments

We thank Katie O’Hearn, MSc (Children’s Hospital of Eastern Ontario Research Institute) for methodological assistance, Douglas M. Salzwedel, MLIS (Therapeutics Initiative, University of British Columbia) for developing and executing the electronic search strategies; Healthy Active Living and Obesity (HALO) research students for their work in level 1 review and data extraction; the team from New York University who conducted the data quality assurance; and David Stein, Carmen H.J. Lim, and Kate H. Jun for critically reviewing the manuscript as external reviewers.

## Funding

No funding

### Data sharing statement

All analyses conducted in this study are fully reproducible. The study-level data will be made publicly available on the Open Science Framework (OSF) shortly following the publication of this pre-print version and will be accessible at https://osf.io/r37mu. Concurrently, the complete analytic codebase used the meta-analysis, and the interactive web application will be released in a single GitHub repository https://github.com/MarcusVVLopes/smu_meta_repo. The live application, deployed a https://marcuslopes.shinyapps.io/smu_meta/, allows for real-time visualization of study-level and pooled estimates under alternative sensitivity analysis criteria to enhance transparency and facilitate knowledge translation.

### CRediT authorship contribution statement

**MVVL:** Conceptualization, Methodology, Validation, Formal analysis, Investigation, Data curation, Writing-Original Draft, Visualization, Project administration; **KB:** Methodology, Data curation, Project administration, Writing - Review & Editing; **AD**: Methodology, Data curation, Writing - Review & Editing;**AG**: Methodology, Data curation, Writing - Review & Editing; **NG**: Data curation, Writing - Review & Editing; **AA**: Data curation, Writing - Review & Editing; **JL**: Data curation, Writing - Review & Editing; **JH**: Conceptualization, Writing - Review & Editing; **ZR**: Conceptualization, Writing - Review & Editing; **JPC**: Writing - Review & Editing, Supervision.; **GSG**: Conceptualization, Methodology, Writing - Review & Editing, Supervision.

