## Appendix for "Effect of Social Media Constraints on Mental Health: A Systematic Review and Meta-Analysis of Experiments"

**eMethods 1**
**PRISMA Checklist**

| Section and Topic | Item # | Checklist item | Location where item is reported |
|---|---|---|---|
| **TITLE** | | | |
| Title | 1 | Identify the report as a systematic review. | p. 1 |
| **ABSTRACT** | | | |
| Abstract | 2 | See the PRISMA 2020 for Abstracts checklist. | p. 2 |
| **INTRODUCTION** | | | |
| Rationale | 3 | Describe the rationale for the review in the context of existing knowledge. | p. 3 |
| Objectives | 4 | Provide an explicit statement of the objective(s) or question(s) the review addresses. | p. 4 |
| **METHODS** | | | |
| Eligibility criteria | 5 | Specify the inclusion and exclusion criteria for the review and how studies were grouped for the syntheses. | p. 4 |
| Information sources | 6 | Specify all databases, registers, websites, organisations, reference lists and other sources searched or consulted to identify studies. Specify the date when each source was last searched or consulted. | p. 4 |
| Search strategy | 7 | Present the full search strategies for all databases, registers and websites, including any filters and limits used. | Appendix, eMethods 3 |
| Selection process | 8 | Specify the methods used to decide whether a study met the inclusion criteria of the review, including how many reviewers screened each record and each report retrieved, whether they worked independently, and if applicable, details of automation tools used in the process. | p. 4 |
| Data collection process | 9 | Specify the methods used to collect data from reports, including how many reviewers collected data from each report, whether they worked independently, any processes for obtaining or confirming data from study investigators, and if applicable, details of automation tools used in the process. | p. 4 |
| Data items | 10a | List and define all outcomes for which data were sought. Specify whether all results that were compatible with each outcome domain in each study were sought (e.g. for all measures, time points, analyses), and if not, the methods used to decide which results to collect. | PROSPERO registration |
| | 10b | List and define all other variables for which data were sought (e.g. participant and intervention characteristics, funding sources). Describe any assumptions made about any missing or unclear information. | PROSPERO registration |
| Study risk of bias assessment | 11 | Specify the methods used to assess risk of bias in the included studies, including details of the tool(s) used, how many reviewers assessed each study and whether they worked independently, and if applicable, details of automation tools used in the process. | p. 4-5 |
| Effect measures | 12 | Specify for each outcome the effect measure(s) (e.g. risk ratio, mean difference) used in the synthesis or presentation of results. | p. 5 |
| Synthesis methods | 13a | Describe the processes used to decide which studies were eligible for each synthesis (e.g. tabulating the study intervention characteristics and comparing against the planned groups for each synthesis (item #5)). | p. 4-5, Appendix eMethods 3 |

| Section and Topic | Item # | Checklist item | Location where item is reported |
|---|---|---|---|
| | 13b | Describe any methods required to prepare the data for presentation or synthesis, such as handling of missing summary statistics, or data conversions. | p. 5, Appendix eMethods 5 |
| | 13c | Describe any methods used to tabulate or visually display results of individual studies and syntheses. | p. 6 |
| | 13d | Describe any methods used to synthesize results and provide a rationale for the choice(s). If meta-analysis was performed, describe the model(s), method(s) to identify the presence and extent of statistical heterogeneity, and software package(s) used. | p. 6 |
| | 13e | Describe any methods used to explore possible causes of heterogeneity among study results (e.g. subgroup analysis, meta-regression). | p. 6 |
| | 13f | Describe any sensitivity analyses conducted to assess robustness of the synthesized results. | p. 7 |
| Reporting bias assessment | 14 | Describe any methods used to assess risk of bias due to missing results in a synthesis (arising from reporting biases). | p. 5 |
| Certainty assessment | 15 | Describe any methods used to assess certainty (or confidence) in the body of evidence for an outcome. | p. 7 |
| **RESULTS** | | | |
| Study selection | 16a | Describe the results of the search and selection process, from the number of records identified in the search to the number of studies included in the review, ideally using a flow diagram. | p. 8, Figure 1 |
| | 16b | Cite studies that might appear to meet the inclusion criteria, but which were excluded, and explain why they were excluded. | eResults 1 |
| Study characteristics | 17 | Cite each included study and present its characteristics. | eResults 2 |
| Risk of bias in studies | 18 | Present assessments of risk of bias for each included study. | eResults 4 |
| Results of individual studies | 19 | For all outcomes, present, for each study: (a) summary statistics for each group (where appropriate) and (b) an effect estimate and its precision (e.g. confidence/credible interval), ideally using structured tables or plots. | Table 1 |
| Results of syntheses | 20a | For each synthesis, briefly summarise the characteristics and risk of bias among contributing studies. | p. 8 |
| | 20b | Present results of all statistical syntheses conducted. If meta-analysis was done, present for each the summary estimate and its precision (e.g. confidence/credible interval) and measures of statistical heterogeneity. If comparing groups, describe the direction of the effect. | p. 9, Table 1 |
| | 20c | Present results of all investigations of possible causes of heterogeneity among study results. | p. 9 |
| | 20d | Present results of all sensitivity analyses conducted to assess the robustness of the synthesized results. | p. 9-10 |
| Reporting biases | 21 | Present assessments of risk of bias due to missing results (arising from reporting biases) for each synthesis assessed. | eResults 4 |
| Certainty of evidence | 22 | Present assessments of certainty (or confidence) in the body of evidence for each outcome assessed. | eResults 4 |

| Section and Topic | Item # | Checklist item | Location where item is reported |
|---|---|---|---|
| **DISCUSSION** | | | |
| Discussion | 23a | Provide a general interpretation of the results in the context of other evidence. | p. 11-12 |
| | 23b | Discuss any limitations of the evidence included in the review. | p. 13 |
| | 23c | Discuss any limitations of the review processes used. | p. 13 |
| | 23d | Discuss implications of the results for practice, policy, and future research. | p. 13-14 |
| **OTHER INFORMATION** | | | |
| Registration and protocol | 24a | Provide registration information for the review, including register name and registration number, or state that the review was not registered. | p. 1, 4 |
| | 24b | Indicate where the review protocol can be accessed, or state that a protocol was not prepared. | Only the PROSPERO protocol is available |
| | 24c | Describe and explain any amendments to information provided at registration or in the protocol. | eMethods 2 |
| Support | 25 | Describe sources of financial or non-financial support for the review, and the role of the funders or sponsors in the review. | p. 14 |
| Competing interests | 26 | Declare any competing interests of review authors. | p. 14-15 |
| Availability of data, code and other materials | 27 | Report which of the following are publicly available and where they can be found: template data collection forms; data extracted from included studies; data used for all analyses; analytic code; any other materials used in the review. | p. 14 |

*From:* Page MJ, McKenzie JE, Bossuyt PM, Boutron I, Hoffmann TC, Mulrow CD, et al. The PRISMA 2020 statement: an updated guideline for reporting systematic reviews. BMJ 2021;372:n71. doi: 10.1136/bmj.n71. This work is licensed under CC BY 4.0. To view a copy of this license, visit https://creativecommons.org/licenses/by/4.0/

### eMethods 2
### Protocol Deviations

To preserve total methodological transparency, the modifications made to the pre-registered study protocol (PROSPERO CRD42025637996) during the implementation and analysis phases are detailed below:

#### *1. Literature search update*

The registered protocol specified an initial electronic database search executed on November 19, 2024, with records from MEDLINE and Embase restricted to publications from 2024 to minimise overlap with Cochrane CENTRAL. To ensure the review reflected the most current available evidence prior to final synthesis, a formal update search was conducted on October 3, 2025. The temporal restriction for MEDLINE and Embase was consequently extended to include records published in 2024 and 2025.

#### *2. Transition to a multilevel random-effects framework*

The protocol prespecified standard random-effects models with the Hartung-Knapp-Sidik-Jonkman (HKSJ) adjustment for pooling effect sizes, with a fallback to fixed-effects models for outcomes with fewer than five study comparisons. In practice, several studies contributed multiple treatment arms or reported multiple outcomes within the same domain, introducing statistical dependencies that a standard two-level model cannot adequately accommodate. Accordingly, a multilevel random-effects framework was adopted, with random intercepts nested at the study and sample levels. Statistical inference was based on t-distribution tests with containment degrees of freedom, which provide more appropriate small-sample-adjusted uncertainty estimates for multilevel structures than the originally planned unilevel HKSJ procedure.

#### *3. Formalisation of the effect size estimation pipeline*

The protocol broadly stated that mean pairwise differences in change scores or post-intervention end-differences would serve as the effect metric, without specifying algebraic estimators or procedures for handling variance adjustments across heterogeneous trial designs. To improve precision and standardise calculations across studies, the Morris (2008) dppc2 estimator was adopted as the primary metric for pre-test/post-test control group designs. This required study-specific pre-post correlation coefficients (r), which primary studies routinely omit. A hierarchical imputation procedure was therefore added: missing correlations were first imputed using domain-specific means estimated from studies with available individual participant data; where domain-level estimation was not possible, a conservative fallback of $r = 0.60$ – the lowest average observed across outcomes – was applied. An F-statistic-to-SMD conversion formula was additionally introduced to recover effect size estimates from studies with insufficient summary statistics for direct standardised calculations.

#### *4. Omission of planned moderation analyses due to reporting inconsistencies*

The registered protocol prespecified meta-regression analyses examining whether intervention effects varied according to participant-level characteristics (age, gender, specific platform use, degree of social media use, and prior mental health conditions) and study-level characteristics (clinical vs. non-clinical samples, intervention length, type of social media manipulation [reduction vs. abstinence], adherence to the intervention, objective vs. subjective measurement of social media use, year of data collection [before or after the predominance of smartphones], and risk-of-bias level)

Because few primary studies reported summary statistics disaggregated by participant-level characteristics, within-study subgroup analyses were not possible. Accordingly, age, gender, and degree of social media use were examined as study-level moderators using aggregate values (i.e., mean age, proportion of female participants, and mean daily social media use at baseline), rather than as participant-level characteristics as originally intended.

Three additional moderators could not be evaluated:

Specific platform use: primary studies rarely isolated effects by individual platform (e.g., Instagram vs. TikTok); most restricted overall social media or use without platform-level disaggregation.

Prior mental health conditions: granular individual-level diagnostic classifications or stratified baseline symptom scores were not available in the study-level aggregate data, preventing formal moderation analysis by clinical status.

Intervention adherence: Quantitative compliance metrics (e.g., verified minutes of social media use reduced per day) were highly non-standardised across primary studies and selectively omitted from published reports, making it impossible to construct a consistent continuous moderator. One study additionally documented a failed manipulation due to non-adherence; however, as only a single such case was identified, a categorical comparison between adherent and non-adherent conditions was not feasible.

Measurement method (objective vs. subjective): this moderator was included in the protocol under the assumption that studies using objective measurement might achieve higher adherence. Given that adherence itself was a prespecified moderator and could not be examined (as described above), the measurement method indicator was excluded to avoid modelling a proxy for an unexaminable construct

***5. Addition of post-hoc sensitivity analyses***

The registered protocol did not prespecify any formal sensitivity analyses. Four sensitivity analyses were conducted post-registration in response to methodological considerations that emerged during data extraction and analysis.

Exclusion of studies classified as problematic cases: studies were flagged as potentially problematic if they presented (1) outlying or highly influential effect sizes, (2) evidence of small-sample bias, (3) documented non-adherence to the social media constraint protocol, or (4) an unclear or unconfirmed absence of manipulation in the control condition. These cases were excluded in a sensitivity analysis to assess the robustness of pooled estimates to studies with evidence of implementation concerns.

Exclusion of single-item outcome measures: outcomes assessed with single-item indicators rather than validated multi-item scales were excluded in a separate sensitivity analysis, given the greater susceptibility of single-item measures to measurement error and their potential to inflate standardised effect sizes.

Exclusion of between-group post-intervention-only designs: analyses were repeated after excluding effect estimates from between-group randomised controlled trials that measured outcomes at post-intervention only, as such designs do not permit estimation of change-score-based effect sizes (dppc1 and dppc2).

Alternative effect size estimator: pooled effects were recalculated using the dppc1 estimator, which applies group-specific rather than pooled baseline standard deviations, to assess the sensitivity of results to the choice of primary estimator. Additionally, post-intervention-only SMDs were computed to replicate the approach used in prior meta-analyses in this area and to directly compare estimates across methodological approaches.

***6. Addition of population-level impact analyses***

The registered protocol specified that objectives were limited to estimating pooled clinical effect sizes and exploring study-level moderators through meta-regression. To translate clinical effect sizes into public health terms, population-level impact analyses were developed post-registration. Pooled Hedges's g values for internalizing symptom domains were converted to odds ratios using the Cox method and combined with multinational prevalence estimates of intense social media use (43%, from the Health Behaviour in School-aged Children survey)[1] to compute Potential Impact Fractions under partial reduction and complete elimination counterfactual scenarios.

**eMethods 3**
**Search Strategy**

The following databases were searched: MEDLINE, Embase, APA PsycINFO, and the Cochrane Central Register of Controlled Trials (CENTRAL). Because MEDLINE and Embase are routinely searched for CENTRAL, results from those databases were restricted to records published in 2024 and 2025. The search strategy was developed and implemented by a librarian with expertise in systematic reviews.

Search Strategy

| Query | Search Terms Used |
|---|---|
| 1 | exp mental disorders/ or mental health/ or (mental* adj3 (condition* or disorder* or distress* or health* or ill*)).tw,kf,kw. |
| 2 | exp anxiety disorders/ or anxiety/ or exp body image/ or depression/ or exp "feeding and eating disorders"/ or exp mood disorders/ or exp stress, psychological/ or exp stress disorders, traumatic/ or exp suicide/ or exp self-injurious behavior/ or ((behav* adj2 (disorder* or health condition* or illness*)) or ((externali* or internali*) adj1 (behavio* or disorder* or problem*)) or (affective disorder* or anorex* or anxiet* or avoidant or binge-eat* or body disatisfaction* or body identity or body image* or bulimi* or depressed or depression or depressive or diabulimi* or disordered eating or eating disorder* or eating syndrom* or emotional disorder* or emotional distress* or emotional health or emotional illness* or feeding disorder* or food addict* or internalis* or internaliz* or loneliness or lonely or loner? or mood? or neuroses or neurosis or neurotic* or orthexia* or panic attack* or panic disorder* or postrauma* or psychiat* or psychologi* condition* or psychologi* distress* or psychologi* health or psychologi* ill* or psychopath* or psychot* or PTSD or rumination syndrom* or stress or stresses or stressful or stressor? or trauma* or suicid* or self-harm* or self-injur* or wellbeing or well-being)).tw,kf,kw. |
| 3 | social media/ or ((facebook* or facetime or Instagram* or instant messag* or live chat* or messaging platform* or messaging software* or skyp* or sms or snapchat* or social messaging or social media or social network* or texting or text messag* or tiktok* or tweets or twitter or whatsapp) adj4 (abstain* or abstinen* or break? or constrain* or curtail* or cut or cutback* or declin* or decreas* or diminish* or expos* or less* or limit* or lower* or manipul* or quit* or ration* or reduc* or shorte* or shrink* or stop* or use* or usage)).tw,kf,kw. |
| 4 | exp randomized controlled trial/ or controlled clinical trial.pt. or (randomi* or assigned or control* or experimental or group or groups or placebo or randomly or trial).ab. or dt.fs. |
| 5 | exp animals/ not humans/ |
| 6 | 4 not 5 |
| 7 | (1 or 2) and 3 and 6 |
| 8 | 7 and 2024*.dt. |
| 9 | exp mental disease/ or mental health/ or (mental* adj3 (condition* or disorder* or distress* or health* or ill*)).tw,kf,kw. |
| 10 | exp anxiety disorder/ or anxiety/ or exp body image/ or exp depression/ or exp eating disorder/ or exp mood disorder/ or exp stress, psychological/ or exp mental stress/ or exp suicidal behavior/ or automutilation/ or ((behav* adj2 (disorder* or health condition* or illness*)) or (affective disorder* or anorex* or anxiet* or avoidant or binge-eat* or body disatisfaction* or body identity or body image* or bulimi* or depressed or depression or depressive or diabulimi* or disordered eating or eating disorder* or eating syndrom* or emotional disorder* or emotional distress* or emotional health or emotional illness* or externalis* or externaliz* or feeding disorder* or food addict* or internalis* or internaliz* or loneliness or lonely or loner? or mood? or neuroses or neurosis or neurotic* or orthexia* or panic attack* or panic disorder* or postrauma* or psychiat* or psychologi* condition* or psychologi* distress* or psychologi* health or psychologi* ill* or psychopath* or psychot* or PTSD or rumination syndrom* or stress or stresses or stressful or stressor? or trauma* or suicid* or self-harm* or self-injur* or wellbeing or well-being)).tw,kf,kw. |

| Query | Search Terms Used |
|---|---|
| 11 | exp social media/ or ((facebook* or facetime or Instagram* or instant messag* or live chat* or messaging platform* or messaging software* or skyp* or sms or snapchat* or social messaging or social media or social network* or texting or text messag* or tiktok* or tweets or twitter or whatsapp) adj4 (abstain* or abstinen* or break? or constrain* or curtail* or cut or cutback* or declin* or decreas* or diminish* or expos* or less* or limit* or lower* or manipul* or quit* or ration* or reduc* or shorte* or shrink* or stop* or use* or usage)).tw,kf,kw. |
| 12 | exp randomized controlled trial/ or controlled clinical trial.pt. or (randomi* or assigned or control* or experimental or group or groups or placebo or randomly or trial).ab. or dt.fs. |
| 13 | (exp animal/ or animal.hw. or nonhuman/) not (exp human/ or human cell/ or (human or humans).ti.) |
| 14 | 12 not 13 |
| 15 | (9 or 10) and 11 and 14 |
| 16 | 15 and 2024*.dc. |
| 17 | exp mental disorders/ or mental health/ or (mental* adj3 (condition* or disorder* or distress* or health* or ill*)).tw,kw. |
| 18 | exp anxiety disorders/ or anxiety/ or exp body image/ or depression/ or exp "feeding and eating disorders"/ or exp mood disorders/ or exp stress, psychological/ or exp stress disorders, traumatic/ or exp suicide/ or exp self-injurious behavior/ or ((behav* adj2 (disorder* or health condition* or illness*)) or (affective disorder* or anorex* or anxiet* or avoidant or binge-eat* or body disatisfaction* or body identity or body image* or bulimi* or depressed or depression or depressive or diabulimi* or disordered eating or eating disorder* or eating syndrom* or emotional disorder* or emotional distress* or emotional health or emotional illness* or externalis* or externaliz* or feeding disorder* or food addict* or internalis* or internaliz* or loneliness or lonely or loner? or mood? or neuroses or neurosis or neurotic* or orthexia* or panic attack* or panic disorder* or postrauma* or psychiat* or psychologi* condition* or psychologi* distress* or psychologi* health or psychologi* ill* or psychopath* or psychot* or PTSD or rumination syndrom* or stress or stresses or stressful or stressor? or trauma* or suicid* or self-harm* or self-injur* or wellbeing or well-being)).tw,kw. |
| 19 | social media/ or ((facebook* or facetime or Instagram* or instant messag* or live chat* or messaging platform* or messaging software* or skyp* or sms or snapchat* or social messaging or social media or social network* or texting or text messag* or tiktok* or tweets or twitter or whatsapp) adj4 (abstain* or abstinen* or break? or constrain* or curtail* or cut or cutback* or declin* or decreas* or diminish* or expos* or less* or limit* or lower* or manipul* or quit* or ration* or reduc* or shorte* or shrink* or stop* or use* or usage)).tw,kw. |
| 20 | (17 or 18) and 19 |
| 21 | exp mental disorders/ or mental health/ or (mental* adj3 (condition* or disorder* or distress* or health* or ill*)).ti,ab,id. |
| 22 | exp anxiety disorders/ or anxiety/ or exp body image/ or depression/ or exp "feeding and eating disorders"/ or exp mood disorders/ or exp stress, psychological/ or exp stress disorders, traumatic/ or exp suicide/ or exp self-injurious behavior/ or ((behav* adj2 (disorder* or health condition* or illness*)) or (affective disorder* or anorex* or anxiet* or avoidant or binge-eat* or body disatisfaction* or body identity or body image* or bulimi* or depressed or depression or depressive or diabulimi* or disordered eating or eating disorder* or eating syndrom* or emotional disorder* or emotional distress* or emotional health or emotional illness* or externalis* or externaliz* or feeding disorder* or food addict* or internalis* or internaliz* or loneliness or lonely or loner? or mood? or neuroses or neurosis or neurotic* or orthexia* or panic attack* or panic disorder* or postrauma* or psychiat* or psychologi* condition* or psychologi* distress* or psychologi* health or psychologi* ill* or psychopath* or psychot* or PTSD or rumination syndrom* or stress or stresses or stressful or stressor? or trauma* or suicid* or self-harm* or self-injur* or wellbeing or well-being)).ti,ab,id. |
| 23 | social media/ or ((facebook* or facetime or Instagram* or instant messag* or live chat* or messaging platform* or messaging software* or skyp* or sms or snapchat* or social messaging or social media or social network* or texting or text messag* or tiktok* or tweets or twitter or whatsapp) adj4 (abstain* or abstinen* or break? or constrain* or curtail* or cut or cutback* or declin* or decreas* or diminish* |

| Query | Search Terms Used |
|---|---|
| | or expos* or less* or limit* or lower* or manipul* or quit* or ration* or reduc* or shorte* or shrink* or stop* or use* or usage)).ti,ab,id. |
| 24 | (randomi* or assigned or control* or experimental or group or groups or placebo or randomly or trial).tw. |
| 25 | (21 or 22) and 23 and 24 |
| 26 | 8 use medall |
| 27 | 16 use emczd |
| 28 | 20 use cctr |
| 29 | 25 use psyh |
| 30 | or/26-29 |
| 31 | remove duplicates from 30 |
| 32 | 31 use medall |
| 33 | 31 use emczd |
| 34 | 31 use cctr |
| 35 | 31 use psyh |

*Note. This table demonstrates the reasons for exclusion of a citation in the second round of reviews. Searches were conducted using an Ovid multi-database search and duplicate records were removed online giving preference to MEDLINE, then Embase, then CENTRAL, with no field preference. Lines 1-8 are optimized for MEDLINE. Lines 9-16 are optimized for Embase, lines 17-20 are optimized for CENTRAL, and lines 21-25 are optimized for PsycINFO. Lines 26-35 isolate the records to the database the search was designed for, combine those sets and then remove duplicate records and finally, isolate the records from each database again so each can be downloaded and imported into the citation manager using a database-specific import filter.*

**eMethods 4**
**Quality Assurance**

Title and abstract screening and full-text review were conducted independently in duplicate by two randomly paired reviewers who underwent training and calibration using a set of 200 records. Reference lists of studies included after full-text review and of relevant previous meta-analyses were screened by AG and AD. Disagreements at any stage were resolved by MVVL, with unresolved cases adjudicated by GSG.

Data extraction was performed independently and in duplicate by two research staff, with conflicting entries resolved by a third reviewer. Remaining uncertainties were referred to MVVL. All study-level variables of interest were prespecified in the registered protocol (PROSPERO: CRD42025637996).

Whenever summary statistics were insufficient for meta-analysis, corresponding authors were contacted and invited to contribute data. Authors were asked to supply summary statistics for the overall sample. Non-responding authors were contacted on up to three occasions over a six-month period. Where authors provided individual participant data in lieu of summary statistics, only the requested summary statistics were computed from those data, with no further use of the participant-level information. Where individual participant data were publicly available in data repositories, summary statistics were re-estimated directly from source data to maximise precision.

Summary statistics derived from individual participant data – whether provided by authors or obtained from public repositories – may differ marginally from those reported in the original publications, as no observations were excluded unless values were deemed implausible based on criteria specified in the original report. Summary statistics extracted from published reports or derived from individual participant data subsequently underwent an independent verification process, in which a second pair of reviewers – not involved in the original extraction – checked all entries against source documents. The data processing pipeline developed by MVVL was additionally reviewed by an independent analyst for accuracy and reproducibility.

### eMethods 5
### Effect Size Calculations

Effect sizes were calculated from study-level means and standard deviations (SD) of outcomes per condition and expressed as standardised mean differences (SMDs) with correction for small samples (Hedge's g). Three estimators were applied depending on study design availability: (a) the Morris (2008) $d_{ppc2}$ estimator as the primary metric for pretest–posttest control group (PPC) designs;[2] (b) the Morris (2008) $d_{ppc1}$ estimator, retained as an alternative for sensitivity analyses;[2] and (c) the post-only SMD for as an alternative for sensitivity analysis and for studies employing post-intervention-only between-groups designs. All three estimators and their sampling variance formulae are derived from Morris (2008) and are described in full below.

For PPC designs, the pre–post correlation coefficient (r) was incorporated into the calculation of sampling variances. When individual participant data were publicly available, study-specific pre-post correlations were calculated empirically for each outcome. For studies lacking this information, the mean correlation estimated across available studies within the same outcome domain was used to impute missing values. When outcome-specific correlations could not be estimated, a conservative value of r = 0 60, corresponding to the lowest average observed across all outcomes, was applied. In cases of insufficient summary statistics for standardised calculations, effect sizes were obtained through F-statistic to SMD conversion formulae.[3]

***$d_{ppc1}$*:** Formally characterized by Morris (2008), the estimator standardizes each group's pre-to-post change by that group's own pretest standard deviation before computing the between-group difference. Because each group's change is normalized against its respective baseline variability, separate pretest SDs appear in the denominator for each group, so $d_{ppc1}$ does not presuppose equality of pretest variances across conditions and is appropriate when baseline variance heterogeneity cannot be ruled out. Each group-specific change is corrected for small-sample bias through separate multiplicative factors $cT$ and $cC$, the Hedges' correction applied within each group independently. The sampling variance of $d_{ppc1}$ accounts for the within-subject pre-post correlation (ρ). Morris (2008) therefore recommends $d_{ppc2}$ over $d_{ppc1}$ when baseline homoscedasticity can be supported.

Effect size formula:

$$d_{ppc1} = \left(\frac{M_{post,T} - M_{pre,T}}{SD_{pre,T}}\right) - \left(\frac{M_{post,C} - M_{pre,C}}{SD_{pre,C}}\right)$$

Variance formula:

$$\sigma^2(d_{ppc1}) = c_T^2\left(\frac{2(1-\rho)}{n_T}\right)\left(\frac{n_T-1}{n_T-3}\right)\left(1+\frac{n_T\delta_T^2}{2(1-\rho)}\right) - \delta_T^2$$
$$+c_C^2\left(\frac{2(1-\rho)}{n_C}\right)\left(\frac{n_C-1}{n_C-3}\right)\left(1+\frac{n_C\delta_C^2}{2(1-\rho)}\right) - \delta_C^2$$

***$d_{ppc2}$*:** The $d_{ppc2}$ estimator which was adopted as the primary metric in the present review, standardizes the total between-group difference in change scores by the pooled pretest standard deviation. This is directly analogous to Cohen's d applied to gain scores, with the advantage that the resulting metric is commensurable with conventional between-group SMDs, facilitating cross-study and cross-design comparability. Unlike $d_{ppc1}$, a single small-sample bias correction factor $c_p$ is applied multiplicatively to the pooled point estimate rather than to each group separately. The sampling variance of $d_{ppc2}$ incorporates a single pooled within-group pre-post correlation (ρ). The pooled pretest SD (SD pre) is calculated as the square root of the pooled within-group pretest variance, weighted by degrees of freedom. Morris (2008) demonstrates that, when the homoscedasticity assumption holds at baseline, $d_{ppc2}$ yields less biased estimates and smaller sampling variances than $d_{ppc1}$, offering higher statistical power and a common effect size scale directly comparable to post-only estimators.

Effect size formula:

$$d_{\text{ppc2}} = c_p \frac{\left(M_{\text{post},T} - M_{\text{pre},T}\right) - \left(M_{\text{post},C} - M_{\text{pre},C}\right)}{SD_{\text{pre}}}$$

where the pooled standard deviation is defined as

$$SD_{pre} = \sqrt{\frac{(n_T - 1)SD^2_{\text{pre},T} + (n_C - 1)SD^2_{\text{pre},C}}{n_T + n_C - 2}}$$

Variance formula:

$$\sigma^2\left(d_{\text{ppc2}}\right) = 2\left(c_p^2\right)(1 - \rho)\left(\frac{n_T + n_C}{n_T n_C}\right)\left(\frac{n_T + n_C - 2}{n_T + n_C - 4}\right)\left(1 + \frac{\Delta^2}{2(1 - \rho)}\left(\frac{n_T + n_C}{n_T n_C}\right)\right) - \Delta^2$$

**Post-SMD:** The post-only SMD ($d_{post}$) was applied to studies employing a post-intervention-only between-groups design in which no pretest measurements were recorded, following the standard Hedges' g formulation. This estimator was also applied to all studies for sensitivity analyses examining the impact of distinct estimators. This estimator calculates the standardized mean difference between treatment and control posttest scores using the pooled posttest standard deviation ($s_p$) as the denominator, equivalent to the conventional two-group SMD described by Borenstein et al. (2009). Because pretest data are absent, no pre-post correlation is incorporated into either the point estimate or the sampling variance. Small-sample bias is corrected by multiplying d post by the factor J (df = n 1 + n 2 − 2) to yield Hedge's g.

Effect size formula:

$$d = \frac{M_1 - M_2}{s_p}$$

Variance formula

$$s_p = \sqrt{\frac{(n_1 - 1)S_1^2 + (n_2 - 1)S_2^2}{n_1 + n_2 - 2}}$$

***Estimated Pre-post Correlations***

| Outcome | Average correlation | Studies [contributed correlation] |
|---|---|---|
| Anxiety Symptoms | 0.73 | Davis et al & Thai et al 2024 [cor = 0.83] |
| | | Allcott et al 2020 [cor = 0.6] |
| | | Maerevoet et al 2025 [cor = 0.77] |
| Body Image | 0.88 | De Hesselle et al 2024 [cor = 0.87] |
| | | Davis et al & Thai et al 2024 [cor = 0.89] |
| Depressive Symptoms | 0.70 | Walsh et al 2024 [cor = 0.79] |
| | | Davis et al & Thai et al 2024 [cor = 0.68] |
| | | Allcott et al 2020 [cor = 0.68] |
| | | Maerevoet et al 2025 [cor = 0.66] |
| Eating Disorders/Pathology | 0.83 | Mikami et al 2024 [cor = 0.83] |
| FoMO/Nomophobia | 0.70 | De Hesselle et al 2024 [cor = 0.65] |
| | | Davis et al & Thai et al 2024 [cor = 0.75] |
| | | Mikami et al 2024 [cor = 0.69] |
| Happiness/Positive Affect | 0.65 | Walsh et al 2024 [cor = 0.61] |
| | | De Hesselle et al 2024 [cor = 0.53] |
| | | Vanman et al 2018 [cor = 0.65] |
| | | Allcott et al 2020 [cor = 0.65] |
| | | Maerevoet et al 2025 [cor = 0.82] |
| Internalizing Symptoms | 0.60 | De Hesselle et al 2024 [cor = 0.6] |
| | | Mikami et al 2024 [cor = 0.61] |
| Life Satisfaction | 0.66 | Walsh et al 2024 [cor = 0.6] |
| | | De Hesselle et al 2024 [cor = 0.69] |
| | | Vanman et al 2018 [cor = 0.67] |
| | | Allcott et al 2020 [cor = 0.66] |
| Loneliness/Isolation | 0.76 | Walsh et al 2024 [cor = 0.74] |
| | | De Hesselle et al 2024 [cor = 0.74] |
| | | Hall et al 2021 [cor = 0.71] |
| | | Vanman et al 2018 [cor = 0.9] |
| | | Mikami et al 2024 [cor = 0.77] |
| | | Allcott et al 2020 [cor = 0.69] |
| Negative Affect/Emotions | 0.76 | Walsh et al 2024 [cor = 0.76] |
| | | Vanman et al 2018 [cor = 0.74] |
| | | Maerevoet et al 2025 [cor = 0.78] |
| Perceived Stress | 0.57 | Walsh et al 2024 [cor = 0.71] |
| | | De Hesselle et al 2024 [cor = 0.46] |
| | | Vanman et al 2018 [cor = 0.44] |
| | | Maerevoet et al 2025 [cor = 0.68] |
| Problematic SM/Internet | 0.61 | De Hesselle et al 2024 [cor = 0.61] |
| Self-Esteem | 0.88 | Walsh et al 2024 [cor = 0.84] |
| | | Maerevoet et al 2025 [cor = 0.93] |
| Well-being | 0.72 | Hall et al 2021 [cor = 0.74] |
| | | Allcott et al 2020 [cor = 0.71] |

***R Implementation***

This R function, compute_es_cor, is designed to calculate effect sizes (Hedges' g) and their corresponding sampling variances for meta-analyses of pre-post control group designs. It specifically implements the formulas described above.

| Argument | Description |
|---|---|
| dat | A data frame containing the study results. |
| measure | Character vector. Choose "ppc1", "ppc2", or "post_only". Multiple can be selected. |
| r_vals | The pre-post correlation coefficient (*r*). Can be a single value or a column in dat. |
| id_cols | Character vector of columns to retain for study identification (e.g., author, year). |
| outcome_col | The column name identifying the specific outcome measured. |
| m_t0, m_t1 | Mean of the treatment group at pre-test (*t0*) and post-test (*t1*). |
| sd_t0, sd_t1 | Standard deviation of the treatment group at pre-test and post-test. |
| n_t | Sample size of the treatment group. |
| m_c0, m_c1 | Mean of the control group at pre-test (*t0*) and post-test (*t1*). |
| sd_c0, sd_c1 | Standard deviation of the control group at pre-test and post-test. |
| n_c | Sample size of the control group. |
| flip_outcomes | Optional vector of outcome names where the scale is reversed (e.g., if a higher score indicates a "worse" result). The function will multiply these by -1. |

Usage example:

```
results <- compute_es_cor(
  dat = my_meta_data,
  measure = "ppc2",
  r_vals = 0.5, # Assuming a constant pre-post correlation of 0.5
  id_cols = c("study_id", "author", "year"),
  m_t0 = treatment_mean_pre,
  m_t1 = treatment_mean_post,
  sd_t0 = treatment_sd_pre,
  n_t = treatment_n,
  m_c0 = control_mean_pre,
  m_c1 = control_mean_post,
  sd_c0 = control_sd_pre,
  n_c = control_n,
  flip_outcomes = c("depression_score", "anxiety_scale")
)
```

The function returns a data frame containing the original identification columns (id_cols); eff_type, the method used (ppc1, ppc2, or post_only); yi, the calculated effect size (Hedges' g); vi, the sampling variance; and cor_pp, the correlation value used in the calculation.

R Function:

```
compute_es_cor <- function(
  dat,
  measure = c("ppc1", "ppc2", "post_only"),
  r_vals = cor_pp_calc,
  id_cols = c("ref_id", "author", "outcome"),
  outcome_col = outcome,
  m_t0 = mean_treat_t0,
  m_t1 = mean_treat_t1,
  sd_t0 = sd_treat_t0,
  sd_t1 = sd_treat_t1,
  n_t = n_treat,
  m_c0 = mean_control_t0,
  m_c1 = mean_control_t1,
  sd_c0 = sd_control_t0,
  sd_c1 = sd_control_t1,
  n_c = n_control,
  flip_outcomes = NULL
) {

  # 1. Dependency Check
  required_pkgs <- c("dplyr", "purrr", "tibble", "metafor")
  missing_pkgs <- required_pkgs[!sapply(required_pkgs, requireNamespace, quietly = TRUE)]
  if (length(missing_pkgs) > 0) {
    stop("The following packages are required: ", paste(missing_pkgs, collapse = ", "))
  }

  measure <- match.arg(measure, several.ok = TRUE)

  # 2. Data Preparation
  dat1 <- dat %>%
    tibble::rowid_to_column(".row_id") %>%
    dplyr::mutate(
      .sign = dplyr::case_when(
        !is.null(flip_outcomes) & (.data[[outcome_col]] %in% flip_outcomes) ~ -1,
        .default = 1
      ),
      r = {{ r_vals }}
    )

  # Hedges' g correction factor J
  calc_j <- function(mi) {
    ifelse(mi <= 1, NA_real_,
           exp(lgamma(mi/2) - log(sqrt(mi/2)) - lgamma((mi - 1)/2)))
  }

  # 3. Calculation Pipeline
  compute_one_measure <- function(m) {
    base <- dat1 %>% dplyr::mutate(.measure = m)

    if (m == "ppc1") {
      # Morris (2008) Eq. 16
      base %>%
        dplyr::mutate(
          d_T = ({{ m_t1 }} - {{ m_t0 }}) / {{ sd_t0 }},
          d_C = ({{ m_c1 }} - {{ m_c0 }}) / {{ sd_c0 }},
          cf_T = calc_j({{ n_t }} - 1),
          cf_C = calc_j({{ n_c }} - 1),
          g_T = cf_T * d_T,
          g_C = cf_C * d_C,
          facT = ({{ n_t }} - 1) / ({{ n_t }} - 3),
          facC = ({{ n_c }} - 1) / ({{ n_c }} - 3),
          var_T = (cf_T^2) * (2 * (1 - r) / {{ n_t }}) * facT * (1 + ({{ n_t }} * d_T^2) / (2 * (1 - r)))
- d_T^2,
          var_C = (cf_C^2) * (2 * (1 - r) / {{ n_c }}) * facC * (1 + ({{ n_c }} * d_C^2) / (2 * (1 - r)))
- d_C^2,
          yi = (g_T - g_C) * .sign,
```

```
        vi = var_T + var_C
      )
    } else if (m == "ppc2") {
      # Morris (2008) Eq. 24
      base %>%
        dplyr::mutate(
          df = {{ n_t }} + {{ n_c }} - 2,
          N = {{ n_t }} + {{ n_c }},
          sd_pool = sqrt((({{ n_t }} - 1) * {{ sd_t0 }}^2 + ({{ n_c }} - 1) * {{ sd_c0 }}^2) / df),
          d = (({{ m_t1 }} - {{ m_t0 }}) - ({{ m_c1 }} - {{ m_c0 }})) / sd_pool,
          cf = calc_j(df),
          yi = (cf * d) * .sign,
          vi = (cf^2) * 2 * (1 - r) * (N / ({{ n_t }} * {{ n_c }})) * ((N - 2) / (N - 4)) * (1 + (d^2 / (2
* (1 - r) * (N / ({{ n_t }} * {{ n_c }}))))) - (d^2)
        )
    } else if (m == "post_only") {
      base %>%
        dplyr::mutate(
          df = {{ n_t }} + {{ n_c }} - 2,
          sd_pool = sqrt((({{ n_t }} - 1) * {{ sd_t1 }}^2 + ({{ n_c }} - 1) * {{ sd_c1 }}^2) / df),
          d = ({{ m_t1 }} - {{ m_c1 }}) / sd_pool,
          cf = calc_j(df),
          yi = (cf * d) * .sign,
          vi = (cf^2) * (1 / {{ n_t }} + 1 / {{ n_c }}) + (yi^2) / (2 * df)
        )
    }
  }

  # 4. Final Formatting
  res <- purrr::map_dfr(measure, compute_one_measure) %>%
    dplyr::mutate(eff_type = .measure) %>%
    dplyr::select(dplyr::all_of(id_cols), eff_type, yi, vi, cor_pp = r)

  return(res)
}
```

**eResults 1**
**Reasons for Exclusion**

| Citation | Reason for exclusion |
|---|---|
| Unknown authors. (2025) The impact of social media detox on mental health: a randomized control trial. | Full text not available |
| Unknown authors. (2025) Reducing Excessive Smartphone Use with Smartphone-enabled Treatment (RESET): a parallel-group, single-blinded, randomized controlled trial. | Full text not available |
| Esmaeili Rad, M., & Ahmadi, F. (2018). A new method to measure and decrease the online social networking addiction. Asia-Pacific psychiatry: official journal of the Pacific Rim College of Psychiatrists, 10(4), e12330. https://doi.org/10.1111/appy.12330 | Insufficient data |
| Gajdics, J., & Jagodics, B. (2022). Mobile Phones in Schools: With or Without you? Comparison of Students' Anxiety Level and Class Engagement After Regular and Mobile-Free School Days. Technology, Knowledge and Learning, 27(4), 1095–1113. https://doi.org/10.1007/s10758-021-09539-w | Insufficient intervention length (less than 24 hours - pre and post-int. within 24hrs) |
| Grobelny, J., Glinka, M., & Chirkowska-Smolak, T. (2024). The impact of hedonic social media use during microbreaks on employee resources recovery. Scientific reports, 14(1), 21603. https://doi.org/10.1038/s41598-024-72825-x | Insufficient intervention length (less than 24 hours - pre and post-int. within 24hrs) |
| Mitev, K., Weinstein, N., Karabeliova, S., Nguyen, T.-v., Law, W., & Przybylski, A. (2021). Social media use only helps, and does not harm, daily interactions and well-being. Technology, Mind, and Behavior, 2(1). https://doi.org/10.1037/tmb0000033 | Insufficient intervention length (less than 24 hours - pre and post-int. within 24hrs) |
| Przybylski, A. K., Nguyen, T. T., Law, W., & Weinstein, N. (2021). Does Taking a Short Break from Social Media Have a Positive Effect on Well-being? Evidence from Three Preregistered Field Experiments. Journal of Technology in Behavioral Science, 6(3), 507–514. https://doi.org/10.1007/s41347-020-00189-w | Insufficient intervention length (less than 24 hours - pre and post-int. within 24hrs) |
| King, D. L., Radunz, M., Galanis, C. R., Quinney, B., & Wade, T. (2024). "Phones off while school's on": Evaluating problematic phone use and the social, wellbeing, and academic effects of banning phones in schools. *Journal of behavioral addictions*, *13*(4), 913–922. https://doi.org/10.1556/2006.2024.00058 | No control group |
| Roberts, T. A., Daniels, E. A., Weaver, J. M., & Zanovitch, L. S. (2022). "Intermission!" A short-term social media fast reduces self-objectification among pre-teen and teen dancers. Body image, 43, 125–133. https://doi.org/10.1016/j.bodyim.2022.08.015 | No control group |
| Upendra, S., & Kaur, J. (2024). Break From Digital Screen Using Digital Detox Program in Nursing Students. Nursing & health sciences, 26(3), e13157. https://doi.org/10.1111/nhs.13157 | No control group |
| Wolf A. Facebook and mental wellbeing: a crossover randomised controlled study [version 1; peer review: 1 approved with reservations, 1 not approved]. F1000Research 2016, 5:1311. https://doi.org/10.12688/f1000research.8835.1 | Pre-print version |

| Citation | Reason for exclusion |
|---|---|
| Grahlher, K., Morgenstern, M., Pietsch, B., Gomes de Matos, E., Rossa, M., Lochbühler, K., Daubmann, A., Thomasius, R., & Arnaud, N. (2024). Mobile App Intervention to Reduce Substance Use, Gambling, and Digital Media Use in Vocational School Students: Exploratory Analysis of the Intervention Arm of a Randomized Controlled Trial. JMIR mHealth and uHealth, 12, e51307. https://doi.org/10.2196/51307 | Wrong comparator (social media manipulated for controls) |
| Brown, L., & Kuss, D. J. (2020). Fear of Missing Out, Mental Wellbeing, and Social Connectedness: A Seven-Day Social Media Abstinence Trial. International Journal of Environmental Research and Public Health, 17(12), 4566. https://doi.org/10.3390/ijerph17124566 | Wrong design (not RCT) |
| Stieger, S., & Lewetz, D. (2018). A Week Without Using Social Media: Results from an Ecological Momentary Intervention Study Using Smartphones. Cyberpsychology, Behavior, and Social Networking, 21(10), 618–624. https://doi.org/10.1089/cyber.2018.0070 | Wrong design (not RCT) |
| Wadsley, M., & Ihssen, N. (2023). Restricting social networking site use for one week produces varied effects on mood but does not increase explicit or implicit desires to use SNSs: Findings from an ecological momentary assessment study. PLOS ONE, 18(11), e0293467. https://doi.org/10.1371/journal.pone.0293467 | Wrong design (not RCT) |
| da Silva Pinho, A., Céspedes Izquierdo, V., Lindström, B., & van den Bos, W. (2024). Youths' sensitivity to social media feedback: A computational account. Science advances, 10(43), eadp8775. https://doi.org/10.1126/sciadv.adp8775 | Wrong design (not RCT) |
| Davis, K. C., Boland, J. K., Fernandez, L. A., & Anderson, J. L. (2023). Give me a break: Do mental health breaks from social networking sites correlate with lower psychopathology? —Preliminary findings. Archives of Psychiatry Research: An International Journal of Psychiatry and Related Sciences, 59(2), 285–294. | Wrong design (not RCT) |
| Tuck, A. B., & Thompson, R. J. (2024). Types of social media use are differentially associated with trait and momentary affect. Emotion (Washington, D.C.), 24(7), 1600–1611. https://doi.org/10.1037/emo0001379 | Wrong design (not RCT) |
| Coulthard, N., & Ogden, J. (2018). The impact of posting selfies and gaining feedback (‘likes’) on the psychological wellbeing of 16–25-year-olds: An experimental study. Cyberpsychology: Journal of Psychosocial Research on Cyberspace, 12(2). https://doi.org/10.5817/CP2018-2-4 | Wrong intervention (not social media reduction) |
| Hou, Y., Xiong, D., Jiang, T., Song, L., & Wang, Q. (2019). Social media addiction: Its impact, mediation, and intervention. Cyberpsychology: Journal of Psychosocial Research on Cyberspace, 13(1). https://doi.org/10.5817/CP2019-1-4 | Wrong intervention (not social media reduction) |
| O’Connell, C. (n.d.). How FOMO (Fear of Missing Out), the Smartphone, and Social Media May Be Affecting University Students in the Middle East. | Wrong intervention (not social media reduction) |
| Olson, J. A., Sandra, D. A., Chmoulevitch, D., et al. (2022). A nudge-based intervention to reduce problematic smartphone use: Randomised Controlled Trial. International Journal of Mental Health Addiction. https:// doi. org/ 10. 1007/ s11469- 022- 00826-w | Wrong intervention (not social media reduction) |
| Evans, C.; King, D.L.; Delfabbro, P.H. Effect of brief gaming abstinence on withdrawal in adolescent at-risk daily gamers: A randomized controlled study. Comput. Hum. Behav. 2018, 88, 70–77. | Wrong intervention (not social media reduction) |

| Citation | Reason for exclusion |
|---|---|
| Brailovskaia, J., Delveaux, J., John, J., Wicker, V., Noveski, A., Kim, S., Schillack, H., & Margraf, J. (2023). Finding the “sweet spot” of smartphone use: Reduction or abstinence to increase well-being and healthy lifestyle?! An experimental intervention study. Journal of Experimental Psychology: Applied, 29(1), 149–161. https://doi.org/10.1037/xap0000430 | Wrong intervention (not social media reduction) |
| Affouneh, S., Mahamid, F. A., Berte, D. Z., Shaqour, A. Z., & Shayeb, M. (2021). The efficacy of a training program for social skills in reducing addictive Internet behaviors among Palestinian university students. *Psicologia, reflexao e critica : revista semestral do Departamento de Psicologia da UFRGS*, 34(1), 19. https://doi.org/10.1186/s41155-021-00185-w | Wrong intervention (not social media reduction) |
| Brockmeier, L. C., Mertens, L., Roitzheim, C., Radtke, T., Dingler, T., & Keller, J. (2025). Effects of an intervention targeting social media app use on well-being outcomes: A randomized controlled trial. Applied psychology. Health and well-being, 17(1), e12646. https://doi.org/10.1111/aphw.12646 | Wrong intervention (not social media reduction) |
| Hussain, Z., Ferreira, R., & Kuss, D. J. (2023). The feasibility of smartphone interventions to decrease problematic use of social networking sites: A randomised controlled trial. Psychiatry Research Communications, 3(3), 100132. https://doi.org/10.1016/j.psycom.2023.100132 | Wrong intervention (not social media reduction) |
| Manwong, M., Lohsoonthorn, V., Booranasuksakul, T., & Chaikoolvatana, A. (2018). Effects of a group activity-based motivational enhancement therapy program on social media addictive behaviors among junior high school students in Thailand: a cluster randomized trial. Psychology research and behavior management, 11, 329–339. https://doi.org/10.2147/PRBM.S168869 | Wrong intervention (not social media reduction) |
| Schmidt-Persson, J., Rasmussen, M. G. B., Sørensen, S. O., Mortensen, S. R., Olesen, L. G., Brage, S., Kristensen, P. L., Bilenberg, N., & Grøntved, A. (2024). Screen Media Use and Mental Health of Children and Adolescents: A Secondary Analysis of a Randomized Clinical Trial. JAMA network open, 7(7), e2419881. https://doi.org/10.1001/jamanetworkopen.2024.19881 | Wrong intervention (not social media reduction) |
| Shafi, R. M. A., Nakonezny, P. A., Miller, K. A., Desai, J., Almorsy, A. G., Ligezka, A. N., Morath, B. A., Romanowicz, M., & Croarkin, P. E. (2021). Altered markers of stress in depressed adolescents after acute social media use. Journal of psychiatric research, 136, 149–156. https://doi.org/10.1016/j.jpsychires.2021.01.055 | Wrong intervention (not social media reduction) |
| Throuvala, M. A., Griffiths, M. D., Rennoldson, M., & Kuss, D. J. (2020). Mind over Matter: Testing the Efficacy of an Online Randomized Controlled Trial to Reduce Distraction from Smartphone Use. International journal of environmental research and public health, 17(13), 4842. https://doi.org/10.3390/ijerph17134842 | Wrong intervention (not social media reduction) |
| Yaman, Z., & Yılmaz, M. (2024). The effect of the programme based on Roy adaptation model on social media addiction, healthy lifestyle and self-esteem of nursing students. International journal of nursing practice, 30(4), e13218. https://doi.org/10.1111/ijn.13218 | Wrong intervention (not social media reduction) |
| Zou, H., Chair, S. Y., Feng, B., Liu, Q., Liu, Y. J., Cheng, Y. X., Luo, D., Wang, X. Q., Chen, W., Huang, L., Xianyu, Y., & Yang, B. X. (2024). A Social Media-Based Mindfulness Psycho-Behavioral Intervention (MCARE) for Patients with Acute Coronary Syndrome: Randomized Controlled Trial. *Journal of medical Internet research*, *26*, e48557. https://doi.org/10.2196/48557 | Wrong intervention (not social media reduction) |

| Citation | Reason for exclusion |
|---|---|
| Turel, O., & R Cavagnaro, D. (2019). Effect of Abstinence from Social Media on Time Perception: Differences between Low- and At-Risk for Social Media "Addiction" Groups. The Psychiatric quarterly, 90(1), 217–227. https://doi.org/10.1007/s11126-018-9614-3 | Wrong outcome (not mental health-related) |
| Verduyn, P., Gugushvili, N., Massar, K., Täht, K., & Kross, E. (2020). Social comparison on social networking sites. Current Opinion in Psychology, 36, 32–37. https://doi.org/10.1016/j.copsyc.2020.04.002 | Wrong publication type (e.g., thesis, editorial) |
| Unknown authors. (2021) The effect of social media on academic performance, mental health and sleep. | Wrong publication type (e.g., thesis, editorial) |
| Unknown authors. (2023) Effect of a Week-long Social Media Abstention on Sustained Attention Functions. | Wrong publication type (e.g., thesis, editorial) |
| Unknown authors. (2023) Change in Social Media Use and Well-being Among College Students Receiving a One-week Exercise or Mindfulness Intervention. | Wrong publication type (e.g., thesis, editorial) |
| Brailovskaia, J., Ströse, F., Schillack, H., & Margraf, J. (2020). Less Facebook use – More well-being and a healthier lifestyle? An experimental intervention study. Computers in Human Behavior, 108, 106332. https://doi.org/10.1016/j.chb.2020.106332 | Wrong publication type (e.g., thesis, editorial) |
| Kleefield, P. (2022). Social media use on smartphones and relations to symptoms of depression and anxiety, Self-Esteem, Sleep and Social Comparison: a Tool for Intervention - ProQuest. https://www.proquest.com/openview/9911853806d1fa1108bfd0f2ef4166d3/1?pq-origsite=gscholar&cbl=18750&diss=y | Wrong publication type (e.g., thesis, editorial) |
| Sagaribay, R. III. (2024). Examining the effects of a week-long social media abstinence intervention, iWeek, on general well-being, mental health, and body image concerns in Latina college students. Dissertation Abstracts International Section A: Humanities and Social Sciences, 85(11-A). | Wrong publication type (e.g., thesis, editorial) |
| Van Timmeren, T. (2022, June 20-22) 'Digital detox': the effect of social media abstinence on usage, habits and wellbeing [Paper presentation]. 7th International Conference on Behavioral Addictions. Nottingham, United Kingdom. | Wrong publication type (e.g., thesis, editorial) |
| Hartanto, A., Kasturiratna, K. T. A. S., Kothari, M., Goh, A. Y. H., Quek, F. Y. X., & Majeed, N. M. (2025). Investigating the effect of full and partial social media abstinence on fear of missing out and well-being outcomes: A daily diary experimental approach. Psychology of Popular Media, 14(4), 572–581. https://doi.org/10.1037/ppm0000583 | Inconsistent data between tables and textual description, leading to opposite conclusions |

**eResults 2**
**Characteristics of the Selected Studies**

| Study | Country | *N* | Age (*M*) | % Female | Type of SMU Restriction | Restriction Length (Weeks) | SMU measure | Outcome(s) |
|---|---|---|---|---|---|---|---|---|
| Agadaullina et al. 2021[4] | Russia | 76 | 19.9 | 70.1 | Abstinence | 2 | Questionnaire | Loneliness (Dynamic of Subjective Experience Scale [DSES]: Social Loneliness Scale, 4 items, subscale); Isolation (Dynamic of Subjective Experience Scale [DSES]: Emotional Loneliness, 4 items, subscale) |
| Allcott et al. 2020[5] | USA | 1639 | 32.58 | 57 | Abstinence | 4 | Other | Anxiety and Depressive Symptoms (European Social Survey Well-Being Module); Happiness/Positive Affect (Subjective Happiness Scale [SHS], 2 items); Life Satisfaction (Satisfaction with Life Scale [SWLS], 3 items); Loneliness/Isolation (Loneliness Scale, 3 items, full scale) |
| Asimovic et al. 2021[6] | Bosnia and Herzegovina | 353 | 29.2 | 64 | Abstinence | 1 | Other | Anxiety Symptoms (single item); Depressive Symptoms (single item); Loneliness/Isolation (single item); Happiness/Positive Affect (single item); Life Satisfaction (Single Item); Fulfillment (single item); Boredom (single item); Well-being (Composite score) |
| Brailovskaia et al. 2020[7] | Germany | 286 | 24.78 | 80 | Reduction | 1, 2 | Questionnaire | Depressive Symptoms (Depression Anxiety Stress Scales 21 [DASS-21], depression subscale); Problematic SM/Internet (Bergen Facebook Addiction Scale [BFAS], 6 items, full scale); Life Satisfaction (Satisfaction with Life Scale [SWLS], 5 items, full scale) |
| Brailovskaia et al. 2022[8] | Germany | 322 | 26.01 | 78.7 | Reduction | 1 | Diary/recall | Depressive Symptoms (Depression Anxiety Stress Scales 21 [DASS-21], depression subscale); Problematic SM/Internet (Bergen Social Media Addiction Scale [BSMAS], 6 items); Life Satisfaction (Satisfaction with Life Scale [SWLS], 5 items, full scale), Happiness/Positive Affect (Subjective Happiness Scale [SHS], 4 items) |

| Study | Country | *N* | Age (*M*) | % Female | Type of SMU Restriction | Restriction Length (Weeks) | SMU measure | Outcome(s) |
|---|---|---|---|---|---|---|---|---|
| Brailovskaia et al. 2024[9] | Germany | 166 | 29.72 | 51.45 | Reduction | 1 | Diary/recall | Problematic SM/Internet (Bergen Social Media Addiction Scale [BSMAS], brief); FoMO/Nomophobia (Fear of Missing Out Scale [FoMO]); Well-being (Positive Mental Health Scale [PMH]); Perceived Stress (Depression Anxiety Stress Scales 21 [DASS-21], stress subscale); Work Satisfaction (Utrecht Work Engagement Scale [UWES], brief), Salutogenic Subjective Working Analyses Questionnaire [SALSA-K], General and Facet-Specific Job Satisfaction Scale [KAFA]) |
| Collis et al. 2022[10] | Denmark | 122 | 22.1 | 50.81 | Reduction | 8 | Objective phone report | Life Satisfaction (Satisfaction with Life Scale [SWLS], 5 items, full scale); Well-being (Short Warwick-Edinburgh Mental Well-being Scale [SWEMWBS], 7 items, subscale) |
| Davis & Goldfield 2024[11] & Thai et al. 2023[12] | Canada | 218 | NR | 76.36 | Reduction | 3 | Objective phone report | Anxiety Symptoms (Generalized Anxiety Disorder Scale, [GAD-7], 7 items, full scale); Depressive Symptoms (Center for Epidemiological Studies: Depression Scale [CES-D], 10 items, subscale); FoMO/Nomophobia (Fear of Missing Out Scale [FoMO], 10 items, full scale); Body Image (Body Esteem Scale for Adults and Adolescents: Appearance [BESAA], 5 items, subscale, Body Esteem Scale for Adults and Adolescents: Weight Esteem [BESAA], 5 items, subscale) |
| De Hesselle et al. 2024[13] | Germany | 86 | 23.33 | 100 | Abstinence | 2 | Objective phone report | Body Image (Multidimensional Body-Self Relations Questionnaire [MBSRQ-AS], Appearance Evaluation subscale, Appearance Orientation subscale, Body Area satisfaction subscale, Overweight Preoccupation subscale), Loneliness (UCLA Loneliness Scale [UCLA-3]); Problematic SM/Internet (Smartphone Addiction Scale - Short Version [SAS-SV], full scale); FoMO/Nomophobia (Fear of Missing Out Scale [FoMO]); Life Satisfaction (Satisfaction with Life Scale [SWLS], 5 |

| Study | Country | *N* | Age (*M*) | % Female | Type of SMU Restriction | Restriction Length (Weeks) | SMU measure | Outcome(s) |
|---|---|---|---|---|---|---|---|---|
| | | | | | | | | items, full scale); Perceived Stress (Perceived Stress Scale [PSS-4]); Well-being (Positive and Negative Affect Schedule [PANAS]) |
| Dondzilo et al. 2024[14] | Australia | 57 | 22.17 | 77.19 | Abstinence | 2 | Objective phone report | Eating Disorders/Pathology (Eating Disorder Examination Questionnaire Short [EDE-QS], 12 items, subscale) |
| Faulhaber et al. 2023[15] | USA | 230 | 22 | 73 | Reduction | 2 | Questionnaire | Anxiety Symptoms (Spielberger State-Trait Anxiety Inventory [STAI], 40 items, full scale); Depressive Symptoms (Epidemiological Studies Depression Scale [CES-D], full scale); FoMO/Nomophobia (Fear of Missing Out Scale [FoMO], 10 items, full scale); Loneliness/Isolation (UCLA Loneliness Scale, 20 items, full scale); Negative Affect/Emotions (Positive and Negative Affect Schedule [PANAS], 5 items, subscale); Happiness/Positive Affect (Positive and Negative Affect Schedule [PANAS], 5 items, subscale) |
| Fioravanti et al. 2019[16] | Italy | 80 | 25.05 | 50 | Abstinence | 1 | Questionnaire | Life Satisfaction (Satisfaction with Life Scale [SWLS], 5 items, Global Life Satisfaction subscale); Negative Affect (Positive and Negative Affect Schedule [PANAS], 10 items, Negative Affect subscale); Positive Affect (Positive and Negative Affect Schedule [PANAS], 10 items, Positive Affect subscale); Body Image (State Appearance Comparison Scale [SACS], full scale) |
| Graham et al. 2021[17] | New Zealand | 124 | 22.46 | 75.8 | Reduction | 1 | Objective phone report | Well-being (Warwick-Edinburgh Mental Well-being Scale [WEMWBS], 14 items, full scale) |

| Study | Country | *N* | Age (*M*) | % Female | Type of SMU Restriction | Restriction Length (Weeks) | SMU measure | Outcome(s) |
|---|---|---|---|---|---|---|---|---|
| Hall et al. 2018[18] & 2021[19] | USA | 49 | 26.8 | 78.5 | Abstinence | 1, 3 | Other | Well-being (Short-Form Health Survey [SF-36], 4 items Well-Being subscale); Loneliness/Isolation (Loneliness Scale, 3 items, full scale) |
| Hanley et al. 2019[20] | Australia | 78 | 30.85 | 55.13 | Abstinence | 1 | Other | Life Satisfaction (Quality of Life Employment and Satisfaction Questionnaire-18 [Q-LES-Q-18], 18 items, adapted scale); Negative Affect (Positive and Negative Affect Schedule [PANAS], 10 items, Negative Affect subscale); Happiness/Positive Affect (Positive and Negative Affect Schedule [PANAS], 10 items, Positive Affect subscale) |
| Hunt et al. 2018[21] | USA | 99 | 19.74 | 76 | Reduction | 3 | Objective phone report | Anxiety Symptoms (Spielberger State-Trait Anxiety Inventory [STAI]); Depressive Symptoms (Beck Depression Inventory [BDI-II], 21 items, full scale); FoMO/Nomophobia (Fear of Missing Out Scale [FoMO], 10 items, full scale); Loneliness/Isolation (UCLA Loneliness Scale [UCLA], 20 items, full scale); Self-Esteem (Rosenberg Self-Esteem Scale [RSES], full scale) |
| Hunt et al. 2021[22] | USA | 61 | 20.11 | 74 | Reduction | 3 | Objective phone report | Anxiety Symptoms (Spielberger State-Trait Anxiety Inventory [STAI]); Depressive Symptoms (Beck Depression Inventory [BDI-II], 21 items, full scale); FoMO/Nomophobia (Fear of Missing Out Scale [FoMO], 10 items, full scale); Loneliness/Isolation (UCLA Loneliness Scale [UCLA], 20 items, full scale); Self-Esteem (Rosenberg Self-Esteem Scale [RSES], full scale) |
| Hunt et al. 2023[23] | USA | 94 | NR | 69 | Reduction | 3 | Objective phone report | Anxiety Symptoms (Spielberger State-Trait Anxiety Inventory [STAI]); Depressive Symptoms (Beck Depression Inventory [BDI-II], 21 items, full scale); FoMO/Nomophobia (Fear of Missing Out Scale [FoMO], 10 items, full scale); Loneliness/Isolation (UCLA Loneliness Scale [UCLA], 20 items, full |

| Study | Country | $N$ | Age ($M$) | % Female | Type of SMU Restriction | Restriction Length (Weeks) | SMU measure | Outcome(s) |
|---|---|---|---|---|---|---|---|---|
| | | | | | | | | scale); Self-Esteem (Rosenberg Self-Esteem Scale [RSES], full scale) |
| Lambert et al. 2022[24] | UK | 140 | 29.6 | 62 | Abstinence | 1 | Objective phone report | Anxiety Symptoms (Generalized Anxiety Disorder-7 Scale [GAD-7], 7 items, full scale); Depressive Symptoms (Patient Health Questionnaire-8 [PHQ-8], 8 items, full scale; Well-being (Warwick-Edinburgh Mental Well-being Scale [WEMWBS], 14 items, full scale) |
| Maerevoet et al. 2025[25] | Belgium | 66 | 21.42 | 85.24 | Reduction | 1, 2 | Objective phone report | Anxiety and Depressive Symptoms (Depression Anxiety Stress Scale 21 [DASS-21], Stress subscale); Negative and Positive Affect (Positive and Negative Affect Schedule [PANAS]); Self-Esteem (Rosenberg Self-Esteem Scale [RSES], 10 items, full scale); Perceived Stress (Perceived Stress Scale [PSS]) |
| Mahalingham et al. 2022[26] | Australia | 93 | 21.94 | 52 | Abstinence | 1 | Objective phone report | Anxiety Symptoms (Depression Anxiety Stress Scales-21 [DASS-21], 7 items, anxiety subscale) |
| Mikami et al. 2024[27] | Canada | 214 | 20.93 | 86.82 | Abstinence | 2, 4, 6 | Objective phone report | Internalizing Symptoms (Depression Anxiety Stress Scale 21 [DASS-21]); Eating Disorders/Pathology (Eating Disorder Inventory-2 [EDI-2], 16 items across multiple domains); FoMO/Nomophobia (Fear of Missing Out Scale [FoMO], 10 items, full scale); Loneliness/Isolation (UCLA Loneliness Scale [UCLA], 20 items, full scale) |
| Mosquera et al. 2019[28] | USA | 151 | 20.59 | 65 | Abstinence | 1 | Other | Depressive Symptoms (OECD Better Life, Depression, single item); Happiness/Positive Affect (OECD Better Life, Feel Happy, single item); Life Satisfaction (OECD Better Life, Overall Satisfaction, single item; Life Worthwhile, single item) |

| Study | Country | $N$ | Age ($M$) | % Female | Type of SMU Restriction | Restriction Length (Weeks) | SMU measure | Outcome(s) |
|---|---|---|---|---|---|---|---|---|
| Reed et al. 2023[29] | UK | 33 | 23.48 | 66 | Reduction | 8 | Objective phone report | Anxiety Symptoms (Hospital Anxiety and Depression Scale [HADS], Anxiety subscale); Depressive Symptoms (Hospital Anxiety and Depression Scale [HADS], Depression subscale); Loneliness/Isolation (UCLA Loneliness Scale [UCLA]); Well-being (The Short Form Health Survey-36 [SF-36]); Problematic SM/Internet (Social Media Addiction Scale – Student Form [SMAS], Addiction subscale) |
| Schwarz et al. 2022[30] | Germany | 185 | 22.28 | 76 | Abstinence | 1 | Diary/recall | Depressive Symptoms (Center for Epidemiologic Studies Depression Scale [CES-D], 20 items, full scale); Well-being (Iowa-Netherlands Comparison Orientation Measure [INCOM], General Mental State, single item); Self-Esteem (Rosenberg Self-Esteem Scale [RSES], 10 items, full scale) |
| Seekis et al. 2025[31] | Australia | 81 | 22.71 | 100 | Reduction, Abstinence | 1 | Questionnaire | Body Image (Self-Objectification Beliefs and Behaviours Scale [SOBBS]; Multidimensional Body-Self Relations Questionnaire [MBSRQ]; Body Appreciation Scale-2 [BAS-2]; Sociocultural Attitudes Towards Appearance Questionnaire [SATAQ-4]); Well-being (World Health Organization Five Item Wellbeing Index [WHO-5]) |
| Smith et al. 2024[32] | Canada | 66 | 19.1 | 100 | Abstinence | 1 | Objective phone report | Body Image (Body Image State Scale [BISS], 6 items, full scale; State Self-Esteem Scale [SSES], Appearance subscale); Self-Esteem (State Self-Esteem Scale [SSES], full scale) |
| Thai et al. 2021[33] | Canada | 37 | NR | 67 | Reduction | 3 | Objective phone report | Anxiety Symptoms (Generalized Anxiety Disorder Scale [GAD-7], 7 items, full scale); Body Image (Body Esteem Scale for Adults and Adolescents [BESAA], Appearance and Weight Esteem subscales); Depressive Symptoms (Center for Epidemiologic Studies Depression Scale Revised [CESD-R], 10 items, subscale) |

| Study | Country | *N* | Age (*M*) | % Female | Type of SMU Restriction | Restriction Length (Weeks) | SMU measure | Outcome(s) |
|---|---|---|---|---|---|---|---|---|
| Tromholt et al. 2016[34] | Denmark | 888 | 34 | 86 | Abstinence | 1 | Diary/recall | Life Satisfaction [single item]; Well-being (Composite of five items from the Center for Epidemiologic Studies Depression Scale [CES-D] and four items from the Positive and Negative Affect Schedule [PANAS]) |
| Turel et al. 2018[35] | USA | 555 | 24.01 | 43 | Abstinence | 1 | Questionnaire | Perceived Stress (Perceived Stress Scale [PSS]) |
| Vally et al. 2019[36] | UAE | 78 | 22.13 | 52.6 | Abstinence | 1 | Questionnaire | Loneliness/Isolation (Social and Emotional Loneliness Scale for Adults – Short Form [SELSA], 6 items, subscale); Negative Affect/Emotions (Positive and Negative Affect Schedule [PANAS], Negative subscale); Perceived Stress [PSS]; Happiness/Positive Affect (Positive and Negative Affect Schedule [PANAS], Positive subscale); Life Satisfaction (Satisfaction with Life Scale [SWLS], 5 items, full scale); Perceived Stress (Perceived Stress Scale [PSS]) |
| Van Wezel et al. 2021[37] | Netherlands | 76 | 20.95 | 64.47 | Reduction | 1 | Objective phone report | FoMO/Nomophobia (Fear of Missing Out Scale [FoMO], 10 items, full scale) Negative Affect/Emotions (Positive and Negative Affect Schedule [PANAS], Negative subscale); Perceived Stress [PSS]; Happiness/Positive Affect (Positive and Negative Affect Schedule [PANAS], Positive subscale) |
| Vanman et al. 2018[38] | Australia | 121 | 22.43 | 63.04 | Abstinence | 0.7 | Other | Loneliness/Isolation (Social and Emotional Loneliness Scale for Adults – Short Form [SELSA]); Life Satisfaction (Satisfaction with Life Scale [SWLS], 5 items, full scale); Negative Affect/Emotions (Positive and Negative Affect Schedule [PANAS], Negative subscale); Perceived Stress [PSS]; Happiness/Positive Affect (Positive and Negative Affect Schedule [PANAS], Positive |

| Study | Country | $N$ | Age ($M$) | % Female | Type of SMU Restriction | Restriction Length (Weeks) | SMU measure | Outcome(s) |
|---|---|---|---|---|---|---|---|---|
| | | | | | | | | subscale) Perceived Stress (Perceived Stress Scale [PSS]) |
| Walsh et al. 2024[39] | USA | 147 | 19.3 | 76.9 | Abstinence | 1 | Objective phone report | Happiness/Positive Affect (Monitoring the Future [MtF], happiness single item); Life Satisfaction (Monitoring the Future [MtF], satisfaction single item); Satisfaction with Life Scale [SWLS], 7 items, full scale); Depressive Symptoms (Bentler Inventory of Depression, full scale); Loneliness/Isolation (Monitoring the Future [MtF], 6 items); Negative Affect/Emotions (Affect-Adjective Scale, Negative Affect subscale); Happiness/Positive Affect (Affect-Adjective Scale, Positive Affect subscale); Self-Esteem (Rosenberg Self-Esteem Scale [RSES], 10 items, full scale); Perceived Stress (Perceived Stress Scale – Short Form [PSS], 4 items, subscale) |
| Zhou et al. 2021[40] | Japan | 65 | 28.79 | 60 | Reduction | 1, 2 | Objective phone report | Life Satisfaction (Satisfaction with Life Scale [SWLS] 5 items, full scale); Work Satisfaction (Minnesota Satisfaction Questionnaire [MSQ]) |

**eResults 3**
**Descriptive Statistics at the Study Level**

| Variable | *N* = 35 |
|---|---|
| Region, *n* (%) | |
| East Asia & Pacific | 7 (20%) |
| Europe & Central Asia | 14 (40%) |
| Middle East & North Africa | 1 (3%) |
| North America | 13 (37%) |
| Sample mean age | |
| Mean (*SD*) | 24.1 (4.1) |
| Median (Q1, Q3) | 22.4 (21.2, 26.4) |
| Min, Max | 19.1, 34.0 |
| Missing | 3 |
| Sample proportion of females | |
| Mean (*SD*) | 70 (15) |
| Median (Q1, Q3) | 70 (60, 79) |
| Min, Max | 43, 100 |
| Sample mean social media use | |
| Mean (*SD*) | 100 (46) |
| Median (Q1, Q3) | 93 (64, 129) |
| Min, Max | 25, 195 |
| Missing | 9 |
| Sample type, *n* (%) | |
| Symptomatic | 3 (9%) |
| General population | 32 (91%) |
| Intervention length (weeks) | |
| Mean (*SD*) | 1.88 (1.75) |
| Median (Q1, Q3) | 1.00 (1.00, 2.00) |
| Min, Max | 0.71, 8.00 |
| Manipulation type, *n* (%) | |
| Abstinence | 19 (54%) |
| Reduction | 15 (43%) |
| Abstinence & Reduction | 1 (3%) |
| Measurement type, *n* (%) | |
| Diary/recall | 4 (11%) |
| Objective phone report | 18 (51%) |

| Variable | $N = 35$ |
|---|---|
| Other | 6 (17%) |
| Questionnaire | 7 (20%) |
| Data collection year | |
| Mean (*SD*) | 2019.54 (2.31) |
| Median (Q1, Q3) | 2020 (2018, 2021) |
| Min, Max | 2015, 2024 |
| Risk of bias, *n* (%) | |
| Low risk | 2 (6%) |
| Some concerns | 26 (74%) |
| High risk | 7 (20%) |

*eResults 4*
*Risk of Bias Assessment*

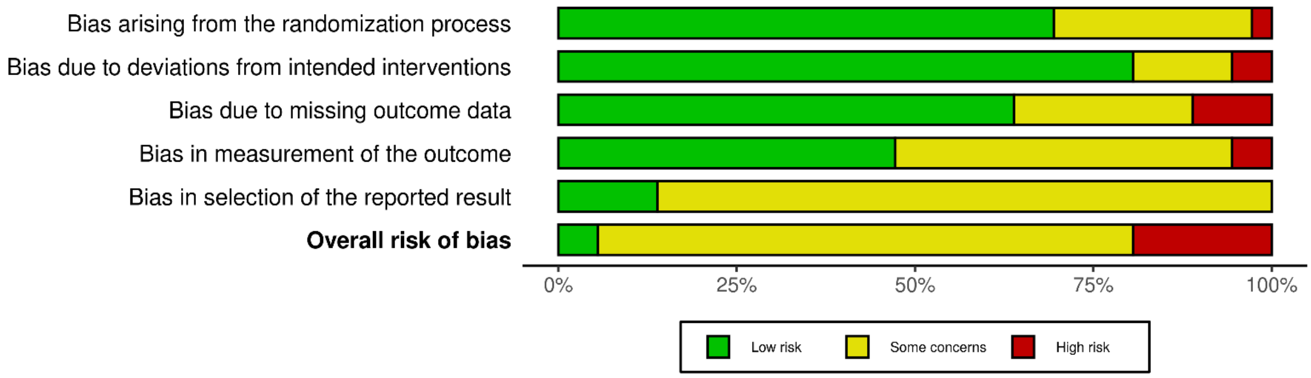

***Risk of Bias Assessment at the Study Level***

Risk of bias domains

| Study | D1 | D2 | D3 | D4 | D5 | Overall |
|---|---|---|---|---|---|---|
| Walsh et al (2024) | + | + | + | + | + | + |
| Dondzilo et al (2024) | - | + | + | + | - | - |
| Brailovskaia et al (2022) | - | + | + | + | - | - |
| Van Wezel et al (2021) | + | + | - | - | - | - |
| Smith et al (2024) | + | + | + | + | - | - |
| Lambert et al (2022) | + | - | X | X | - | X |
| Mahalingham et al (2022) | - | + | + | + | - | - |
| Fioravanti et al (2019) | - | - | X | - | - | X |
| Collis et al (2022) | - | + | + | + | + | - |
| Faulhaber et al (2023) | + | + | + | + | - | - |
| De Hesselle et al (2024) | - | - | - | - | - | - |
| Schwarz et al (2022) | - | X | - | - | - | X |
| Hunt et al (2021) | + | + | + | + | - | - |
| Graham et al (2021) | + | + | + | - | - | - |
| Vally et al (2019) | + | + | + | - | - | - |
| Hunt et al (2018) | + | X | X | - | - | X |
| Tromholt et al (2016) | + | + | + | + | - | - |
| Zhou et al (2021) | - | + | - | + | - | - |
| Asimovic et al (2021) | + | + | X | - | + | X |
| Davis et al (2024) & Thai et al (2023) | + | + | + | + | + | + |
| Thai et al (2021) | + | + | + | - | - | - |
| Brailovskaia et al (2020) | + | + | + | - | - | - |
| Hall et al (2019) & Hall et al (2021) | X | - | + | - | - | X |
| Vanman et al (2018) | + | + | + | + | - | - |
| Mikami et al (2024) | + | + | + | + | - | - |
| Hanley et al (2019) | + | + | - | + | - | - |
| Mosquera et al (2019) | - | + | + | + | - | - |
| Reed et al (2023) | + | + | + | - | - | - |
| Allcott et al (2020) | + | + | - | - | - | - |
| Seekis et al (2025) | + | - | - | - | + | - |
| Turel et al (2018) | + | + | + | - | - | - |
| Brailovskaia et al (2024) | + | + | - | + | - | - |
| Hunt et al (2023) | + | + | + | - | - | - |
| Maerevoet et al (2025) | - | + | + | + | - | - |
| Agadullina et al (2021) | + | + | - | X | - | X |
| Hartanto et al (2025) | + | + | + | - | - | - |

Domains:
D1: Bias arising from the randomization process.
D2: Bias due to deviations from intended intervention.
D3: Bias due to missing outcome data.
D4: Bias in measurement of the outcome.
D5: Bias in selection of the reported result.

Judgement
X High
- Some concerns
+ Low

**eResults 5**
**Studies Flagged as Problematic Cases**

| Outcome | Problematic cases [reason] |
|---|---|
| Depressive Symptoms | Reed et al (2023) [small sample bias]<br>Allcott et al (2020) [problematic design] |
| Perceived Stress | N.A. |
| Anxiety Symptoms | Hunt et al (2018) [outlier effect]<br>Allcott et al (2020) [problematic design] |
| Loneliness/Isolation | Vally et al (2019) [outlier effect]<br>Allcott et al (2020) [problematic design] |
| FoMO/Nomophobia | Van Wezel et al (2021) [problematic design] |
| Negative Affect/Emotions | Van Wezel et al (2021) [problematic design] |
| Well-being | Seekis et al (2025) [outlier effect at 1 week]<br>Allcott et al (2020) [problematic design] |
| Self-Esteem | N.A. |
| Happiness/Positive Affect | Fioravanti et al (2019) [outlier effect]<br>Van Wezel et al (2021) [problematic design]<br>Allcott et al (2020) [problematic design] |
| Life Satisfaction | Vanman et al (2018) [outlier effect]<br>Allcott et al (2020) [problematic design] |
| Body Image | N.A. |

Note: N.A., Not applicable

**eResults 6**

**Sensitivity Analysis: Reanalysis with the exclusion of effects derived from single-item questions**

| Outcome | Main Effects | | | Heterogeneity | | | Publication Bias |
|---|---|---|---|---|---|---|---|
| | *k* | N | Hedges' *g* [95% CI] | *Q* | *p*Q | *I*2 (%) | Egger's *p* |
| Depressive Symptoms | 15 (15) | 2192 | **0.26 [0.12, 0.39]** | 46.013 | 0 | 68.35 | 0.024 |
| Perceived Stress | 7 (7) | 1169 | **0.15 [0.01, 0.29]** | 4.779 | 0.572 | 0 | NA |
| Anxiety Symptoms | 9 (9) | 977 | **0.20 [0.02, 0.37]** | 19.051 | 0.015 | 59.35 | NA |
| Loneliness/Isolation | 13 (16) | 2906 | 0.09 [-0.04, 0.21] | 38.85 | 0.001 | 69.19 | 0.487 |
| FoMO/Nomophobia | 9 (10) | 1214 | **0.14 [0.04, 0.24]** | 5.417 | 0.797 | 0 | 0.411 |
| Negative Affect/Emotions | 8 (8) | 876 | 0.07 [-0.29, 0.43] | 61.872 | 0 | 89.04 | NA |
| Well-being | 10 (11) | 3620 | **0.38 [0.09, 0.67]** | 91.678 | 0 | 93.4 | 0.442 |
| Self-Esteem | 7 (9) | 718 | 0.12 [-0.16, 0.40] | 74.214 | 0 | 87.97 | NA |
| Happiness/Positive Affect | 11 (11) | 2923 | 0.14 [-0.05, 0.32] | 32.555 | 0 | 78.88 | 0.48 |
| Life Satisfaction | 11 (11) | 3027 | 0.11 [-0.08, 0.30] | 33.232 | 0 | 80.41 | 0.422 |
| Body Image | 6 (18) | 569 | 0.26 [0.02, 0.50] | 68.242 | 0.000 | 80.26 | 0.581 |
| Eating Disorders/Pathology | 2 (2) | 244 | 0.15 [-3.01, 3.31] | 6.898 | 0.009 | 85.5 | NA |

*Note: k: Number of studies [number of effect sizes]. N: Total sample size. Hedges' g: Pooled effect size. QE: Q-statistic for heterogeneity. QEp: p-value for Q-statistic. I2: Percentage of total variability due to true effect size differences. Egger's p: p-value for Egger's regression test for publication bias.*

**eResults 7**

**Sensitivity Analysis: Reanalysis with the exclusion of effects derived between-participant designs**

| Outcome | Main Effects | | | Heterogeneity | | | Publication Bias |
|---|---|---|---|---|---|---|---|
| | *k* | N | Hedges' *g* [95% CI] | *Q* | *p*Q | *I*2 (%) | Egger's *p* |
| Depressive Symptoms | 17 (17) | 3982 | **0.23 [0.12, 0.35]** | 54.532 | 0 | 68.85 | 0.014 |
| Perceived Stress | 6 (6) | 1091 | **0.16 [0.01, 0.32]** | 3.637 | 0.603 | 0 | NA |
| Anxiety Symptoms | 10 (10) | 2616 | **0.17 [0.02, 0.32]** | 26.023 | 0.002 | 66.23 | 0.439 |
| Loneliness/Isolation | 12 (15) | 2828 | **0.11 [0.01, 0.22]** | 29.502 | 0.009 | 55.42 | 0.062 |
| FoMO/Nomophobia | 9 (10) | 1214 | **0.14 [0.04, 0.24]** | 5.417 | 0.797 | 0 | 0.411 |
| Negative Affect/Emotions | 7 (7) | 798 | 0.14 [-0.22, 0.51] | 53.269 | 0 | 88.28 | NA |
| Well-being | 9 (10) | 2564 | **0.39 [0.06, 0.72]** | 88.052 | 0 | 92.79 | 0.471 |
| Self-Esteem | 7 (9) | 718 | 0.12 [-0.16, 0.40] | 74.214 | 0 | 87.97 | NA |
| Happiness/Positive Affect | 11 (12) | 2996 | 0.14 [-0.03, 0.30] | 32.842 | 0.001 | 75.05 | 0.198 |
| Life Satisfaction | 11 (13) | 3100 | 0.08 [-0.09, 0.24] | 31.741 | 0.002 | 76.43 | 0.649 |
| Body Image | 6 (18) | 569 | 0.26 [0.02, 0.50] | 68.242 | 0.000 | 80.26 | 0.581 |
| Eating Disorders/Pathology | 2 (2) | 244 | 0.15 [-3.01, 3.31] | 6.898 | 0.009 | 85.5 | NA |

*Note: k: Number of studies [number of effect sizes]. N: Total sample size. Hedges' g: Pooled effect size. QE: Q-statistic for heterogeneity. QEp: p-value for Q-statistic. I2: Percentage of total variability due to true effect size differences. Egger's p: p-value for Egger's regression test for publication bias.*

**eResults 8**

**Sensitivity Analysis: Reanalysis with effects derived from the $d_{PPC1}$ estimator**

| Outcome | Main Effects | | | Heterogeneity | | | Publication Bias |
|---|---|---|---|---|---|---|---|
| | *k* | N | Hedges' *g* [95% CI] | *Q* | *p*Q | *I*2 (%) | Egger's *p* |
| Depressive Symptoms | 18 (18) | 4335 | **0.22 [0.11, 0.32]** | 52.861 | 0 | 62.67 | 0.009 |
| Perceived Stress | 7 (7) | 1169 | 0.13 [-0.02, 0.27] | 4.403 | 0.622 | 0 | NA |
| Anxiety Symptoms | 11 (11) | 2969 | **0.20 [0.07, 0.34]** | 26.874 | 0.003 | 59.97 | 0.503 |
| Loneliness/Isolation | 14 (18) | 3259 | 0.10 [-0.01, 0.20] | 39.991 | 0.001 | 59.55 | 0.539 |
| FoMO/Nomophobia | 9 (10) | 1214 | **0.15 [0.04, 0.26]** | 4.932 | 0.84 | 0 | 0.597 |
| Negative Affect/Emotions | 8 (8) | 876 | 0.06 [-0.27, 0.40] | 54.087 | 0 | 86.4 | NA |
| Well-being | 11 (12) | 3805 | **0.37 [0.11, 0.64]** | 82.849 | 0 | 92.55 | 0.293 |
| Self-Esteem | 7 (9) | 718 | 0.14 [-0.13, 0.41] | 57.002 | 0 | 85.81 | NA |
| Happiness/Positive Affect | 13 (14) | 3427 | 0.12 [-0.01, 0.25] | 30.248 | 0.004 | 63.86 | 0.305 |
| Life Satisfaction | 14 (16) | 4419 | 0.11 [-0.03, 0.24] | 41.066 | 0 | 74.54 | 0.643 |
| Body Image | 6 (18) | 569 | **0.27 [0.02, 0.53]** | 65.669 | 0.000 | 81.22 | 0.542 |
| Eating Disorders/Pathology | 2 (2) | 244 | 0.12 [-2.67, 2.92] | 5.117 | 0.024 | 80.46 | NA |

*Note: k: Number of studies [number of effect sizes]. N: Total sample size. Hedges' g: Pooled effect size. QE: Q-statistic for heterogeneity. QEp: p-value for Q-statistic. I2: Percentage of total variability due to true effect size differences. Egger's p: p-value for Egger's regression test for publication bias.*

**eResults 9**
**Sensitivity Analysis: Reanalysis with effects derived from the post-only SMD estimator**

| Outcome | Main Effects | | | Heterogeneity | | | Publication Bias |
|---|---|---|---|---|---|---|---|
| | *k* | N | Hedges' *g* [95% CI] | *Q* | *p*Q | *I*2 (%) | Egger's *p* |
| Depressive Symptoms | 18 (18) | 4335 | 0.21 [0.10, 0.33] | 40.908 | 0.001 | 54.62 | 0.064 |
| Perceived Stress | 7 (7) | 1169 | 0.15 [-0.13, 0.43] | 15.245 | 0.018 | 64.49 | NA |
| Anxiety Symptoms | 11 (11) | 2969 | 0.24 [0.08, 0.40] | 21.99 | 0.015 | 55.11 | 0.327 |
| Loneliness/Isolation | 14 (16) | 3259 | 0.07 [-0.02, 0.17] | 24.831 | 0.052 | 17.63 | 0.609 |
| FoMO/Nomophobia | 9 (10) | 1214 | 0.22 [0.09, 0.34] | 7.597 | 0.575 | 0 | 0.339 |
| Negative Affect/Emotions | 8 (8) | 876 | 0.11 [-0.14, 0.36] | 17.561 | 0.014 | 56.54 | NA |
| Well-being | 11 (12) | 3805 | 0.32 [0.02, 0.62] | 79.212 | 0 | 92.21 | 0.797 |
| Self-Esteem | 7 (9) | 718 | 0.20 [-0.07, 0.46] | 15.196 | 0.055 | 50.43 | NA |
| Happiness/Positive Affect | 13 (14) | 3427 | 0.10 [-0.02, 0.23] | 22.871 | 0.043 | 49.97 | 0.289 |
| Life Satisfaction | 14 (16) | 4419 | 0.03 [-0.09, 0.16] | 35.887 | 0.002 | 62.16 | 0.296 |
| Body Image | 6 (18) | 569 | 0.16 [-0.11, 0.43] | 24.820 | 0.099 | 50.08 | 0.499 |
| Eating Disorders/Pathology | 2 (2) | 244 | 0.14 [-5.08, 5.35] | 7.269 | 0.007 | 86.24 | NA |

*Note: k: Number of studies [number of effect sizes]. N: Total sample size. Hedges' g: Pooled effect size. QE: Q-statistic for heterogeneity. QEp: p-value for Q-statistic. I2: Percentage of total variability due to true effect size differences. Egger's p: p-value for Egger's regression test for publication bias.*

eResults 10. Study-level moderators examined through meta-regression. Problematic cases removed.

| Moderators | Depressive Symptoms | | Anxiety Symptoms | | Loneliness/Isolation | | FoMO/Nomophobia | | Well-being | | Happiness/Positive Affect | | Life Satisfaction | | Body Image | |
|---|---|---|---|---|---|---|---|---|---|---|---|---|---|---|---|---|
| | *g* [95% CI] | *p* | *g* [95% CI] | *p* | *g* [95% CI] | *p* | *g* [95% CI] | *p* | *g* [95% CI] | *p* | *g* [95% CI] | *p* | *g* [95% CI] | *p* | *g* [95% CI] | *p* |
| **Region** | | 0.17 | | 0.669 | | 0.995 | | 0.409 | | 0.956 | | 0.695 | | 0.775 | | 0.374 |
| East Asia & Pacific | — | | — | | — | | — | | 0.32 [-0.25, 0.88] | | 0.00 [-0.41, 0.42] | | 0.06 [-0.34, 0.45] | | — | |
| Europe & Central Asia | 0.14 [0.01, 0.28] | | 0.26 [0.03, 0.48] | | 0.16 [-0.02, 0.34] | | 0.22 [0.01, 0.43] | | 0.33 [0.04, 0.63] | | 0.16 [-0.10, 0.43] | | 0.17 [-0.00, 0.33] | | 0.11 [-0.29, 0.51] | |
| North America | 0.27 [0.14, 0.40] | | 0.31 [0.10, 0.53] | | 0.16 [0.02, 0.30] | | 0.13 [0.00, 0.25] | | — | | 0.05 [-0.24, 0.34] | | 0.08 [-0.20, 0.37] | | 0.33 [0.00, 0.66] | |
| **Mean age** | 0.02 [-0.09, 0.14] | 0.674 | 0.06 [-0.13, 0.25] | 0.47 | 0.01 [-0.11, 0.13] | 0.922 | 0.08 [-0.05, 0.22] | 0.183 | 0.11 [-0.12, 0.34] | 0.297 | -0.04 [-0.19, 0.12] | 0.611 | 0.02 [-0.11, 0.14] | 0.764 | -0.14 [-0.32, 0.05] | 0.140 |
| **Proportion of females** | -0.07 [-0.18, 0.04] | 0.209 | -0.03 [-0.17, 0.12] | 0.678 | -0.04 [-0.15, 0.07] | 0.457 | -0.05 [-0.17, 0.08] | 0.404 | -0.03 [-0.28, 0.22] | 0.801 | 0.11 [-0.05, 0.28] | 0.144 | 0.00 [-0.13, 0.13] | 0.978 | 0.03 [-0.15, 0.21] | 0.712 |
| **Social media use at baseline** | 0.08 [-0.03, 0.19] | 0.151 | 0.09 [-0.05, 0.22] | 0.165 | -0.04 [-0.18, 0.11] | 0.572 | 0.05 [-0.08, 0.18] | 0.401 | 0.14 [-0.64, 0.93] | 0.599 | 0.13 [-0.00, 0.27] | 0.053 | 0.09 [-0.08, 0.26] | 0.247 | -0.06 [-0.21, 0.10] | 0.329 |
| **Sample type** | | 0.246 | | 0.723 | | — | | 0.808 | | — | | — | | — | | 0.543 |
| Clinical sample | 0.33 [0.09, 0.57] | | 0.31 [0.00, 0.62] | | — | | 0.13 [-0.05, 0.32] | | — | | — | | — | | 0.17 [-0.19, 0.54] | |
| Community sample | 0.18 [0.08, 0.29] | | 0.25 [0.08, 0.42] | | — | | 0.16 [0.03, 0.29] | | — | | — | | — | | 0.30 [0.05, 0.56] | |
| **Intervention length (weeks)** | 0.05 [-0.04, 0.15] | 0.257 | 0.04 [-0.17, 0.24] | 0.691 | 0.10 [0.01, 0.20] | 0.04 | -0.05 [-0.16, 0.05] | 0.273 | -0.12 [-0.37, 0.13] | 0.288 | 0.09 [-0.03, 0.21] | 0.128 | -0.05 [-0.17, 0.07] | 0.398 | -0.11 [-0.28, 0.05] | 0.168 |
| **Manipulation type** | | 0.445 | | 0.873 | | 0.08 | | 0.532 | | 0.724 | | 0.285 | | 0.52 | | 0.784 |
| Abstinence | 0.17 [0.03, 0.31] | | 0.25 [0.02, 0.48] | | 0.07 [-0.03, 0.18] | | 0.10 [-0.12, 0.31] | | 0.33 [0.03, 0.63] | | 0.03 [-0.12, 0.19] | | 0.17 [0.02, 0.33] | | 0.25 [0.03, 0.47] | |
| Reduction | 0.24 [0.11, 0.37] | | 0.27 [0.09, 0.45] | | 0.24 [0.08, 0.40] | | 0.17 [0.04, 0.29] | | 0.25 [-0.13, 0.64] | | 0.16 [-0.04, 0.37] | | 0.09 [-0.12, 0.31] | | 0.27 [0.05, 0.50] | |
| **Study date** | 0.05 [-0.04, 0.15] | 0.267 | -0.04 [-0.18, 0.10] | 0.482 | -0.01 [-0.12, 0.10] | 0.866 | 0.09 [-0.05, 0.22] | 0.166 | 0.04 [-0.21, 0.28] | 0.726 | 0.12 [-0.01, 0.26] | 0.063 | 0.03 [-0.09, 0.16] | 0.601 | 0.05 [-0.12, 0.22] | 0.520 |
| **Risk of Bias** | | 0.704 | | 0.774 | | 0.322 | | — | | 0.708 | | — | | 0.461 | | — |
| Low risk | 0.12 [-0.14, 0.38] | | — | | — | | — | | — | | — | | | | — | |
| Some concerns | 0.23 [0.10, 0.37] | | 0.26 [0.04, 0.48] | | 0.11 [-0.02, 0.24] | | — | | 0.27 [-0.04, 0.58] | | — | | 0.13 [-0.04, 0.29] | | — | |
| High risk | 0.21 [0.01, 0.41] | | 0.31 [-0.01, 0.62] | | 0.21 [0.04, 0.39] | | — | | 0.35 [-0.02, 0.73] | | — | | 0.26 [-0.09, 0.61] | | — | |

Note: g: Hedges' g (Pooled Effect Size for continuous moderators; Estimated Marginal Mean for categorical levels). p: p-value (Omnibus Q-test for categorical headers; Slope coefficient test for continuous variables). CI: Confidence Interval. FoMO: fear of missing out.
